# Immunogenicity of mRNA-1273 and BNT162b2 in Immunocompromised Patients: Systematic Review and Meta-Analysis Using GRADE

**DOI:** 10.1101/2023.08.09.23293898

**Authors:** Sushma Kavikondala, Katrin Haeussler, Xuan Wang, Anne Spellman, Mary T. Bausch-Jurken, Pawana Sharma, Mohammadreza Amiri, Anna Krivelyova, Sonam Vats, Maria Nassim, Nitendra Kumar, Nicolas Van de Velde

## Abstract

**Aim:** Immunocompromised (IC) patients mount poor immune responses to vaccination. Higher-dose COVID-19 vaccines may offer increased immunogenicity.

**Materials & methods:** A pairwise meta-analysis of 98 studies reporting comparisons of mRNA-1273 (50 or 100 mcg/dose) and BNT162b2 (30 mcg/dose) in IC adults was performed. Outcomes were seroconversion, total and neutralizing antibody titers, and cellular immune responses.

**Results:** mRNA-1273 was associated with a significantly higher seroconversion likelihood (relative risk, 1.11 [95% CI, 1.08, 1.14]; *P*<0.0001; *I*^2^*=*66.8%) and higher total antibody titers (relative increase, 50.45% [95% CI, 34.63%, 66.28%]; *P*<0.0001; *I^2^*=89.5%) versus BNT162b2. mRNA-1273 elicited higher but statistically nonsignificant relative increases in neutralizing antibody titers and cellular immune responses versus BNT162b2.

**Conclusion:** Higher-dose mRNA-1273 had increased immunogenicity versus BNT162b2 in IC patients.

## Introduction

Several vaccines against SARS-CoV-2, which causes COVID-19, were developed in response to the COVID-19 pandemic [1]. Two vaccines using novel messenger ribonucleic acid (mRNA) technology were approved for use against COVID-19 [1]: mRNA-1273 (Spikevax^®^, Moderna, Inc., Cambridge, MA, USA) [2] and BNT162b2 (Comirnaty^®^, Pfizer/BioNTech, New York, NY, USA/Mainz, Germany) [3]. Both mRNA-1273 and BNT162b2 administered in a 2-dose series significantly reduced symptomatic infections and hospitalizations in immunocompetent populations evaluated in pivotal studies [4, 5]. The mRNA-1273 and BNT162b2 2-dose primary series were also shown to elicit high neutralizing antibody titers against the spike protein of SARS-CoV-2, as well as high rates of seroconversion in the general population [6, 7].

People who are immunocompromised (IC) generally mount poor immune responses to vaccination because of their immunocompromising conditions or therapies used to treat their underlying diseases, rendering them susceptible to infections [8]. IC populations include but are not limited to patients with cancer, autoimmune diseases, HIV, or primary or secondary immune system deficiencies; solid organ transplant recipients; and patients receiving immunosuppressive therapies (eg, B-cell−depleting agents such as anti-CD20 monoclonal antibodies) [9].

Approximately 10 million people in the United States are considered to be IC [10]; however, people with IC conditions were excluded from participating in phase 2/3 clinical trials of mRNA-1273 and BNT162b2 [4, 5]. Observational studies have demonstrated that IC populations are at increased risk of COVID-19−related morbidity and mortality compared with the general population [8, 11-13]. One study showed that nearly half (44%) of COVID-19−associated hospitalizations among vaccinated people (ie, breakthrough hospitalizations) occurred in IC individuals [14]. IC patients are also at higher risk of longer courses of infection [15-22] and viral evolution [15-18, 20, 23, 24]. Poor humoral immune responses also exacerbate risks posed by new SARS-CoV-2 variants [25-31].

High-dose influenza vaccines have been shown to elicit greater immune responses compared with standard-dose vaccines in IC populations [32-37]. Higher-dose mRNA COVID-19 vaccines may offer similar benefits. In addition to differences in the lipid nanoparticle component of the vaccines, the primary series of mRNA-1273 contains over 3 times the amount of mRNA compared with BNT162b2 (100 mcg/dose vs 30 mcg/dose) and the booster nearly 2 times (50 mcg/dose vs 30 mcg/dose) [2, 3, 38, 39]. Humoral immune responses to COVID-19 vaccines have been shown to correlate with clinical efficacy or effectiveness in general [40-42] and IC populations [43, 44]. Although randomized controlled trials (RCTs) evaluating the comparative immunogenicity of mRNA COVID-19 vaccines in IC populations are lacking, observational studies suggest that there are differences in the immune responses elicited by the mRNA COVID-19 vaccines in IC populations [28, 45], which may affect protection from severe COVID-19 in these vulnerable populations.

National immunization technical advisory groups, including the Advisory Committee on Immunization Practices (ACIP) in the United States, make recommendations on the best use of available vaccines in specific populations [46]. ACIP evaluates available evidence according to the Grading of Recommendations, Assessment, Development and Evaluations (GRADE) framework to formulate vaccine recommendations [47, 48]. For example, a systematic literature review (SLR) and pairwise meta-analysis was performed by ACIP to evaluate the evidence for high-dose influenza vaccines compared with standard-dose influenza vaccines in adults ≥65 years old [49]. The results were evaluated using the GRADE framework and ultimately supported a recommendation for high-dose influenza vaccines in adults ≥65 years old [49, 50].

Determining which of the mRNA-based COVID-19 vaccines offer the highest protection from disease is essential in preventing poor COVID-19−related outcomes in highly susceptible IC populations. Data on the comparative immunogenicity of mRNA-1273 and BNT162b2 in IC populations are therefore urgently needed to inform public health policy. In this SLR and pairwise meta-analysis, we compared seroconversion rates, total and neutralizing anti-spike antibody titers, and cellular immunity levels in IC individuals after vaccination with 2 or 3 doses of mRNA-1273 or BNT162b2. We also applied the GRADE framework to address the following healthcare question: Does the 2-dose mRNA-1273 COVID-19 vaccine primary series (100 mcg mRNA/dose) or primary series and booster (50 or 100 mcg mRNA/dose) have greater immunogenicity in IC populations compared with the 2-dose BNT162b2 COVID-19 vaccine primary series or primary series and booster (30 mcg mRNA/dose irrespective of dose type)?

## Methods

The SLR was performed per the Preferred Reporting Items for Systematic Reviews and Meta-Analyses 2020 framework [51]. A separate meta-analysis evaluating the comparative clinical effectiveness of mRNA-1273 and BNT162b2 was performed using data extracted from the same SLR and published separately [52].

### Search Strategy

As previously described [52], the main search was conducted in the World Health Organization COVID-19 Research Database on April 14, 2022, and updated on December 19, 2022. **Table S1** lists the search strings.

### Study Selection Criteria

Titles and abstracts were screened against inclusion criteria by 2 independent reviewers in level 1. Full texts were evaluated against selection criteria in level 2. A third reviewer arbitrated conflicts between reviewers.

A summary of the population, intervention/exposure, comparison, and outcomes described below is shown in **Table S2**. Studies were included if they were clinical trials, observational studies, or any real-world evidence published as manuscripts, letters, commentaries, abstracts, or posters reporting immunogenicity outcomes in IC individuals ≥18 years of age vaccinated with a homologous primary series of mRNA-1273 or BNT162b2 or the primary series followed by a homologous booster within the same study. We defined the primary series as either 2 doses of mRNA-1273 (100 mcg mRNA/dose) or BNT162b2 (30 mcg mRNA/dose) or used the definition of primary series reported on a per-study basis due to globally varying recommendations for a third vaccine dose in IC populations over time. The booster dose was defined as a homologous third dose of mRNA-1273 (50 or 100 mcg mRNA/dose) or BNT162b2 (30 mcg mRNA/dose). People belonging to clinically extremely vulnerable (CEV) groups 1 or 2 [9], which included transplant recipients, patients with cancer, primary immunodeficiencies, dialysis or severe kidney disease, poorly controlled HIV infection, or autoimmune diseases requiring immunosuppressive therapy, were considered to be IC. Recently published SLRs on the same topic were cross-checked to ensure relevant articles were not omitted. Studies reporting outcomes in pregnant women, current or former smokers, or physically inactive people and those with a heterologous vaccination schedule (ie, mix of mRNA-1273 and BNT162b2), only safety data, or study protocols or economic models were excluded.

### Outcomes

Immunogenicity outcomes were selected based on immune correlates of protection against SARS-CoV-2 investigated in preclinical and clinical studies of COVID-19 vaccines [42, 53, 54]. These correlates of protection are associated with lower risk of severe infection at the population level; however, the threshold of protection for total antibody and neutralizing antibodies has yet to be established [40, 54]. Regulatory authorities have authorized mRNA COVID-19 vaccines for emergency use among different age groups, additional primary series doses for IC populations, and booster doses through immunobridging (ie, predicting vaccine effectiveness based on immunogenicity demonstrated in new populations); the correlates of immunity used were seroresponse rates and neutralizing antibody titers [55-57].

The primary outcome was the percentage of patients achieving seroconversion after vaccination with 2 or 3 doses of mRNA-1273 compared with BNT162b2. Seroconversion was defined as the presence of SARS-CoV-2 anti-spike antibodies above the cutoff value indicated by the specifications of the manufacturer of the assay used to measure antibody titers following vaccination, or as defined in each study based on an evaluation of correlation with plaque reduction neutralization tests. Details on assays used in each study are provided in **Table S3.** Secondary outcomes were total anti-spike binding antibody or immunoglobulin G (IgG) titers (**Table S4**), neutralizing anti-spike antibody titers (**Table S5**), and cellular immune response based on interferon (IFN)-γ or interleukin (IL) levels or CD4^+^/CD8^+^ T-cell levels (**Table S6**).

Outcomes reported after the homologous 2-dose primary series were included in the analysis. If not reported, outcomes following the homologous third dose (booster or third full dose) were included. For studies reporting multiple time points, outcomes assessed 2 weeks after the second dose of the primary series (if only 2 doses were given) or the third dose administered were preferentially included. If outcomes 2 weeks after the last dose administered were not reported, outcomes assessed at time points ≥2 weeks after the final dose were considered instead. For studies reporting multiple IC populations, data were included from populations with the highest number of patients, events, or measured antibody titers for each study. Total patient population and event numbers or rates were required to be reported for seroconversion, and the mean or median was required to be reported for total and neutralizing anti-spike antibody titers. If the total population size per vaccine arm was not reported, the study was excluded from the meta-analysis. Whenever available, antibody titers assessed by the Roche Elecsys platform and in COVID-19–naïve populations were included in the meta-analysis.

Total binding antibody titers were preferentially used for the total anti-spike antibody outcome if reported; otherwise, anti-spike IgG titers were used. Data for the cellular immune response outcome were preferentially taken from IFN-γ− or IL-producing T-cell assays or concentrations, followed by CD4^+^/CD8^+^ T-cell counts or mean spot-forming units from ELISpot assays. If available, composite scores of the data described above were considered first, followed by IFN-γ levels and other IL levels. Studies were required to report mean or median levels. Distribution statistics were imputed from available data if they were not reported [58].

### Data Extraction and Quality Assessment

Publication details, study and patient characteristics, vaccine type and vaccination status, IC condition, background anti-CD20 monoclonal antibody treatment, and immunogenicity data were extracted. Risk of bias (RoB) was assessed in accordance with Cochrane review guidelines [59] using version 2 of the RoB 2 tool [60] for randomized studies and the Newcastle-Ottawa scale [61] for observational studies. Observational studies with <7 and ≥7 stars were considered to have serious and no serious RoB, respectively. Evidence was evaluated based on the GRADE framework [47, 48].

### Statistical Analysis

Irrespective of assay type, random-effects meta-analysis models were used to pool risk ratios (RRs) across studies and absolute effects as risk difference (RD) per 100,000 individuals were calculated across studies for the seroconversion outcome. Inverse variance weights were calculated for individual studies with the DerSimonian-Laird method [62]. Relative increase and corresponding standard errors in total anti-spike binding or IgG antibody titers, neutralizing antibody titers, and cellular immune response outcomes were calculated to compare mRNA-1273 versus BNT162b2 following the Dubey method as suggested in the Cochrane Handbook [58, 63]. Chi-square testing to evaluate heterogeneity across studies was performed [64]. The *I*^2^ statistic was estimated (0%−100%) and interpreted as follows: 0% to 40%, no evidence of heterogeneity or heterogeneity might not be important; >40% to 60%, evidence of moderate heterogeneity; >60% to 75%, evidence of substantial heterogeneity; and >75%, evidence of considerable heterogeneity.

Outcomes were analyzed separately for RCTs and nonrandomized studies. To reduce heterogeneity introduced by differences in the underlying IC condition and therapies used to treat the IC condition, 2 subgroup analyses were conducted. The first subgroup analysis analyzed patients with autoimmune disease, solid organ transplant recipients, patients with solid tumors, and patients with hematologic malignancies separately. In the second subgroup analysis, outcomes were further analyzed separately by prior anti-CD20 treatment versus no treatment in IC patients overall and among patients with autoimmune disease and hematologic malignancies.

## Results

### Overview of Included Studies

Of the 5745 nonduplicate articles identified in the main searches, 130 articles reporting immunogenicity outcomes in IC patients vaccinated with mRNA-1273 versus BNT162b2 in the same study were included in the SLR. Of these, 98 articles were included in the pairwise meta-analysis (**Figure 1**). Reasons for excluding articles from the meta-analysis were that the data were not suitable for meta-analysis (eg, only event rates reported or data not reported by vaccine arm separately; n=13), patients received either a single dose of vaccine or a heterologous vaccine series (n=9), the population did not meet CEV groups 1 and 2 or age inclusion criteria (n=7), and duplicate studies (eg, preprint version of another included study; n=3).

**Figure 1.**
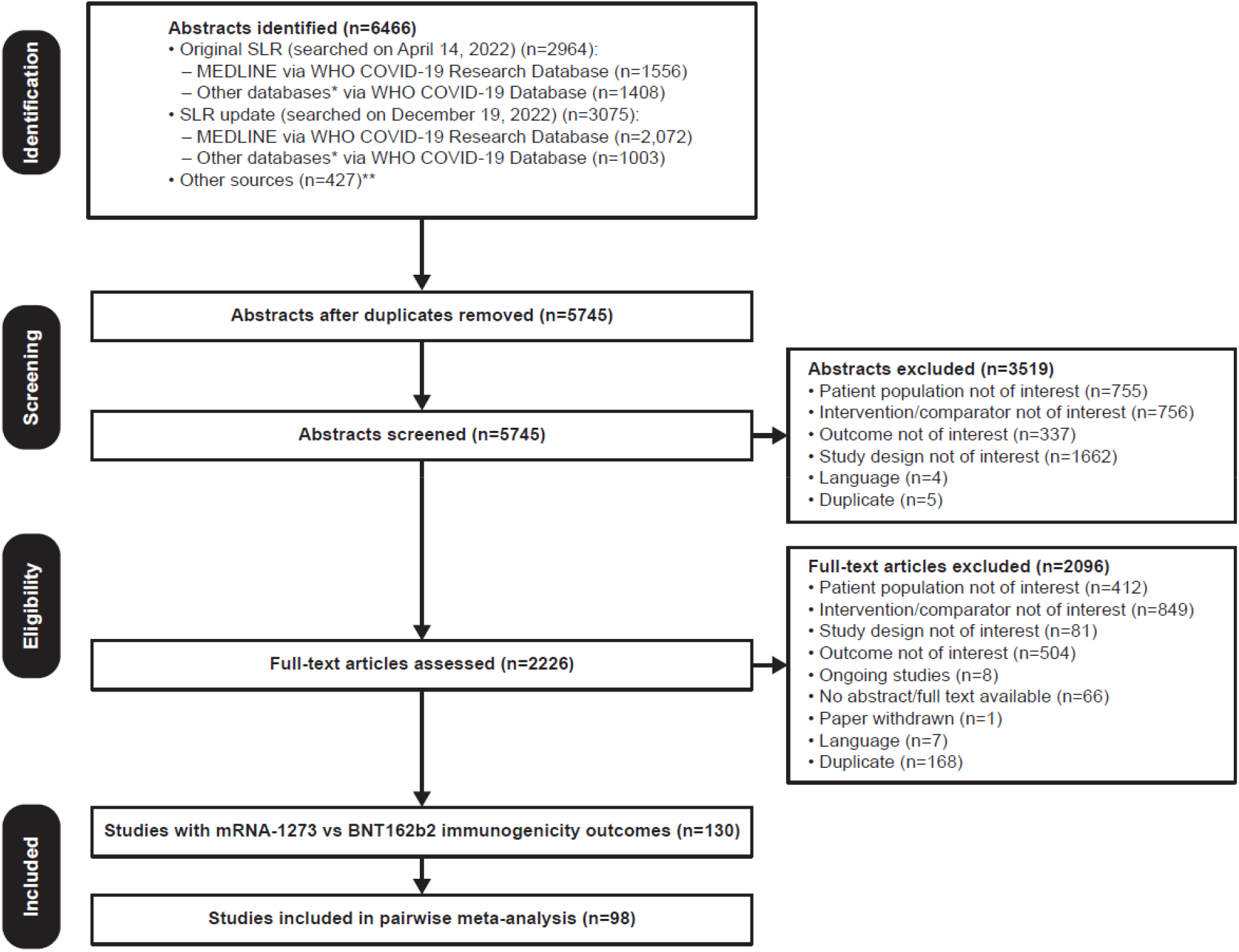
PRISMA flow diagram. Searches were first performed on April 14, 2022, followed by an update on December 19, 2022. *Databases searched include ICTRP, Embase, EuropePMC, medRxiv, Web of Science, ProQuest Central, Academic Search Complete, Scopus, and COVIDWHO. **Includes internal documents from Moderna and recently published SLRs. ICTRP, International Clinical Trials Registry Platform; PRISMA, Preferred Reporting Items for Systematic Reviews and Meta-Analyses; SLR, systematic literature review; WHO, World Health Organization.

**Figure 2.**
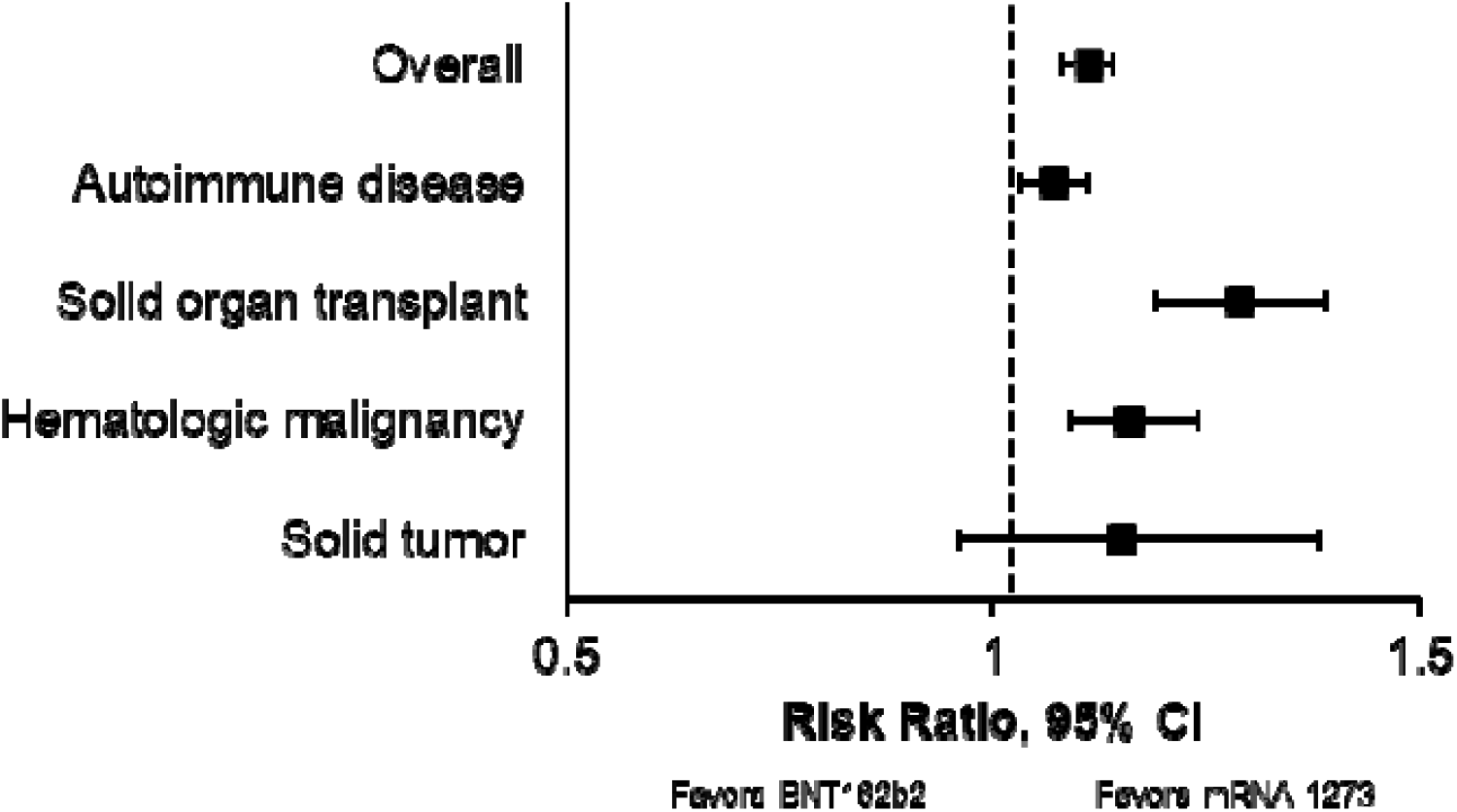
Summary of the seroconversion meta-analysis by disease subgroup. Likelihood of seroconversion in IC patients vaccinated with mRNA-1273 vs BNT162b2 overall and by disease subgroup are shown. IC, immunocompromised.

Characteristics of the studies included in the meta-analysis by outcome are provided in **Tables S3**−**S6.** Of the 98 articles included in the pairwise meta-analysis overall, 1 study was an RCT [65] and the remaining 97 were nonrandomized studies. Seventy-nine nonrandomized studies comprising 23,135 patients vaccinated with mRNA-1273 (n=9664) or BNT162b2 (n=13,471) were analyzed for the overall seroconversion outcome. Forty-five nonrandomized studies comprising 8913 patients vaccinated with mRNA-1273 (n=3038) or BNT162b2 (n=5875) and 1 RCT comprising 50 patients vaccinated with mRNA-1273 (n=24) or BNT162b2 (n=26) were analyzed for the overall total anti-spike binding antibody or IgG titer outcome. Seven nonrandomized studies comprising 592 patients vaccinated with mRNA-1273 (n=236) or BNT162b2 (n=356) were analyzed for the overall neutralizing anti-spike antibody titer outcome. Fourteen nonrandomized studies comprising 2583 patients vaccinated with mRNA-1273 (n=878) or BNT162b2 (n=1705) were analyzed for the overall cellular immune response outcome. Most studies evaluated the 2-dose primary series (n=81, 82.7%). Patients received 2 doses of the primary series followed by 1 booster dose in 17 studies (17.3%) [66-82]. Most studies did not specify whether the mRNA-1273 booster dose was a true booster (ie, 50 mcg mRNA/dose) or a third dose of the primary series (ie, 100 mcg mRNA/dose).

Assessment of RoB was performed; nearly half of the studies (n=47, 48.0%) were determined to have no serious RoB (**Table S7; Table S8**). Forty-five studies (45.9%) had serious RoB, largely because non-IC cohorts were not included or not described if included, only 1 cohort was included, or cohort comparability was not assessed or described if multiple cohorts were included. RoB was not assessed for 6 articles (6.1%) because they were abstracts or research letters.

### Seroconversion

The meta-analysis of 79 nonrandomized studies overall found that seroconversion was significantly more likely with mRNA-1273 compared with BNT162b2 in IC patients (RR, 1.11 [95% CI, 1.08, 1.14]; *P*<0.0001; **Table 1**). The heterogeneity between studies was considered substantial (*I*^2^=66.8%). Expressed as absolute differences, we found that mRNA-1273 vaccination would result in seroconversion of 8170.28 more IC patients per 100,000 IC patients (95% CI, 6279.09, 10,061.47; *P*<0.0001; I^2^=60.3%) compared with BNT162b2 vaccination.

**Table 1.**
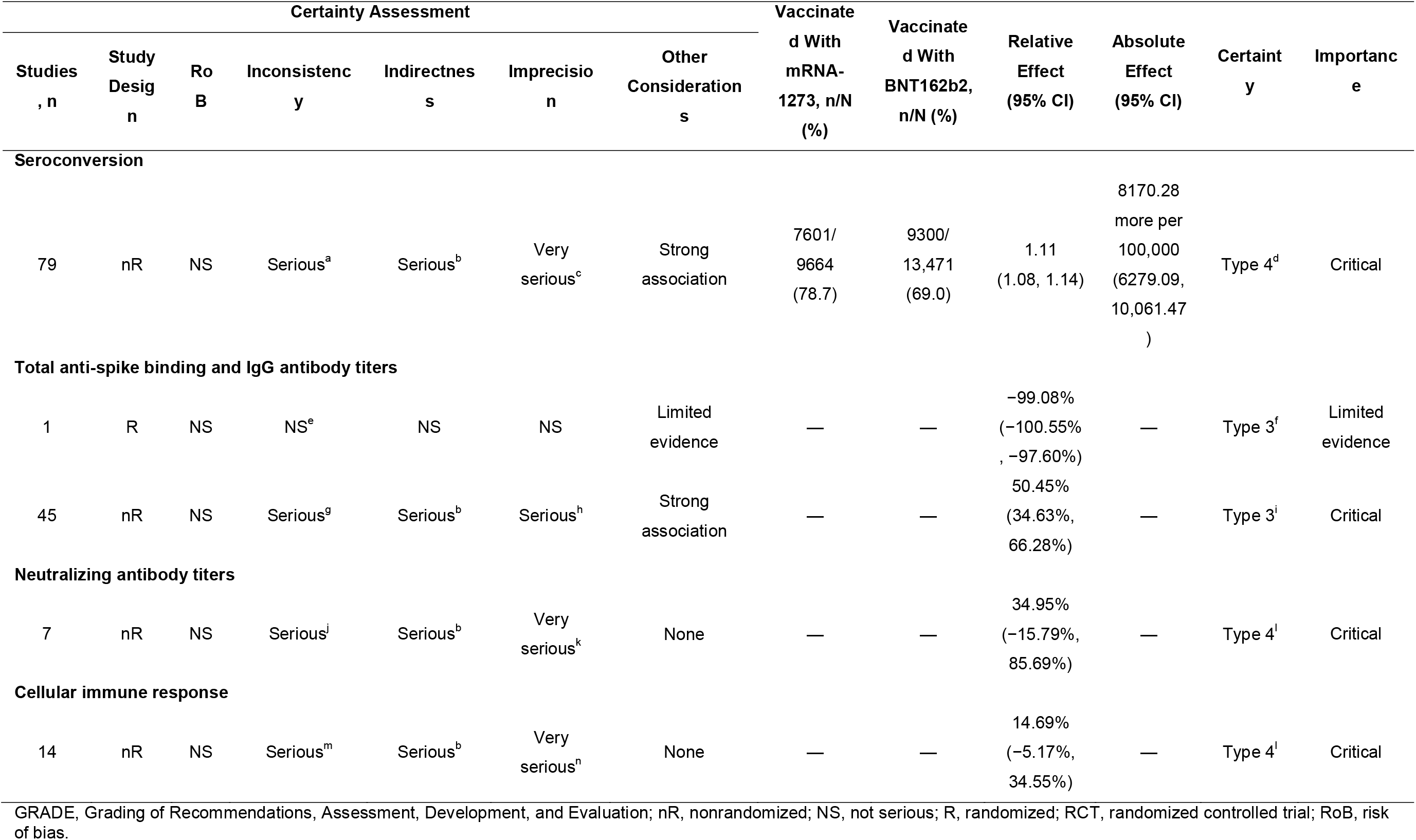
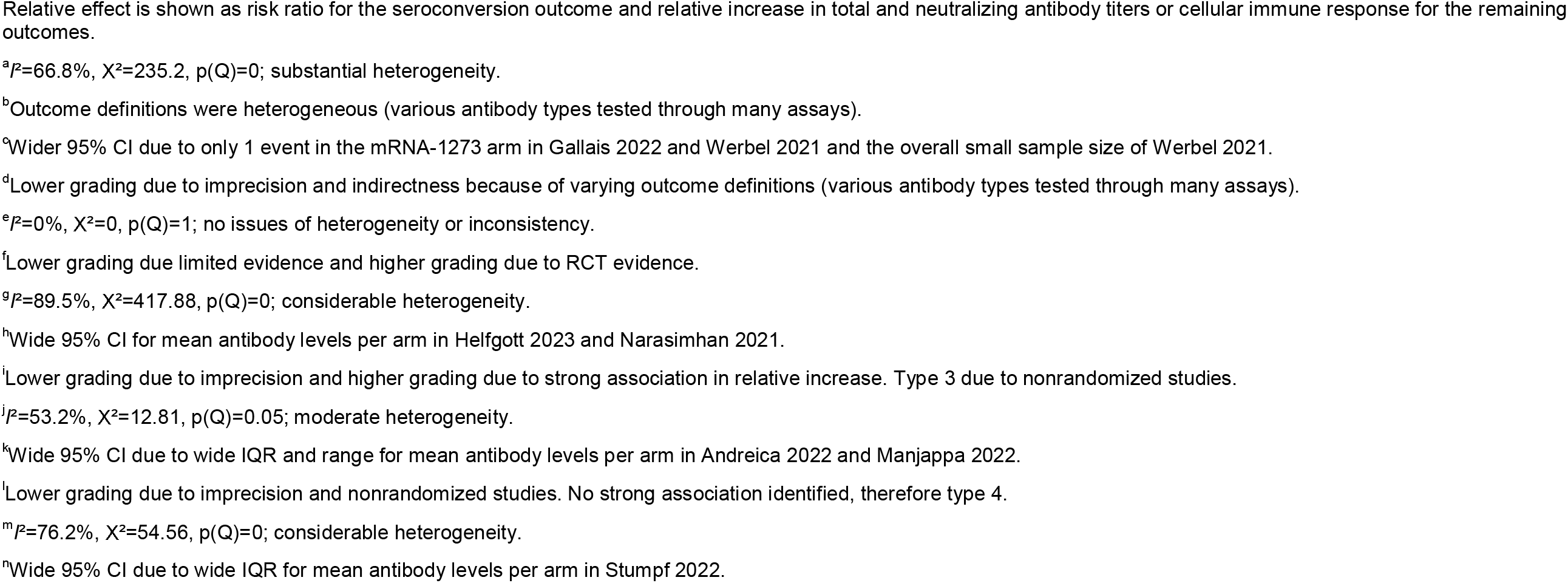
GRADE Summary of Findings Overall.

Similar trends in seroconversion were observed in IC patients analyzed by disease subgroup (**Table 2**). Among solid organ transplant recipients, mRNA-1273 was significantly more likely to result in seroconversion compared with BNT162b2 (RR, 1.29 [95% CI, 1.19, 1.39]; *P*<0.0001; I^2^=43.5%). In patients with autoimmune disease, seroconversion was significantly more likely for mRNA-1273 compared with BNT162b2 (RR, 1.07 [95% CI, 1.03, 1.11]; *P*=0.0002; *I^2^*=58.1%). Patients with hematologic malignancies were also significantly more likely to seroconvert following mRNA-1273 vaccination versus BNT162b2 (RR, 1.16 [95% CI, 1.09, 1.24]; *P*<0.0001; *I*^2^=57.0%). mRNA-1273 was not associated with a statistically significant difference in likelihood of seroconversion compared with BNT162b2 in patients with solid tumors.

**Table 2.**
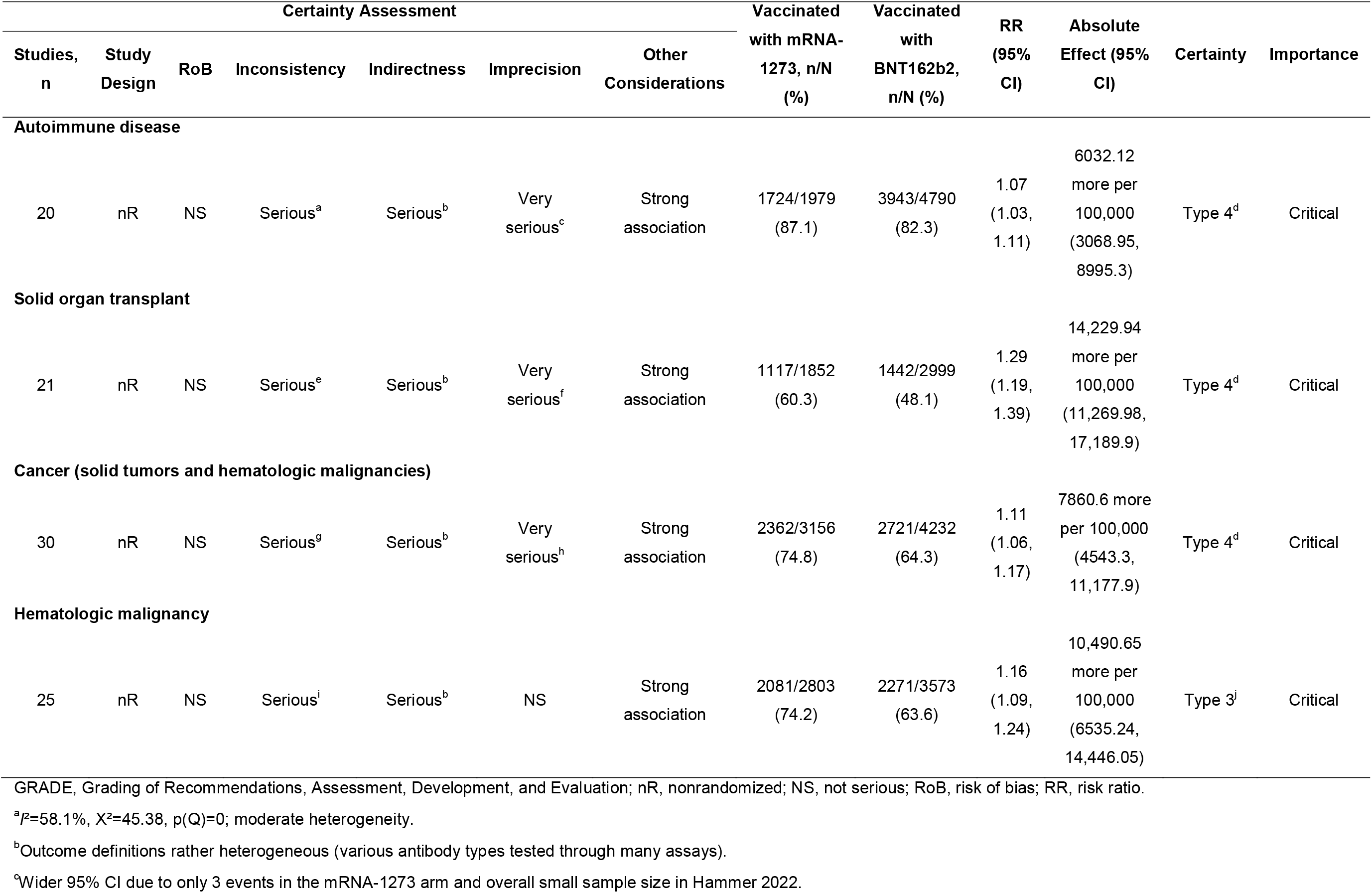
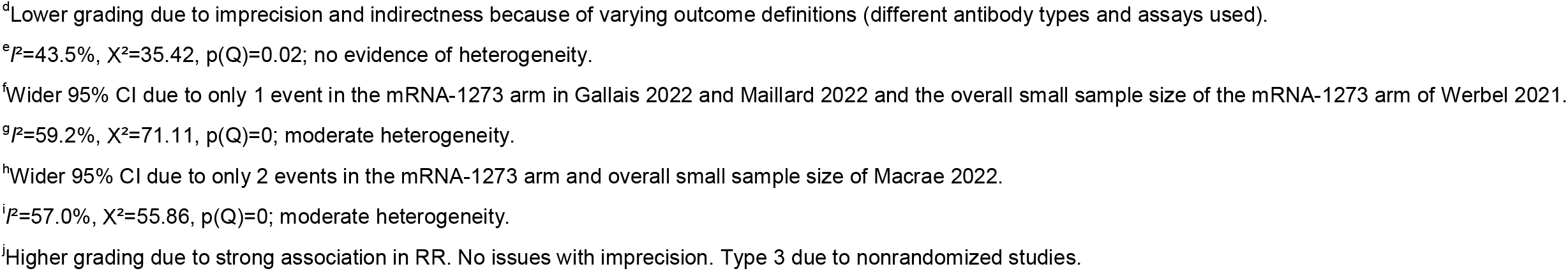
GRADE Summary of Seroconversion by Disease Subgroup.

| Certainty Assessment |  |  |  |  |  |  | Vaccinated<br>with mRNA-1273, n/N<br>(%) | Vaccinated<br>with<br>BNT162b2,<br>n/N (%) | RR<br>(95%<br>CI) | Absolute<br>Effect (95%<br>CI) | Certainty | Importance |
| --- | --- | --- | --- | --- | --- | --- | --- | --- | --- | --- | --- | --- |
| Studies,<br>n | Study<br>Design | RoB | Inconsistency | Indirectness | Imprecision | Other<br>Considerations |  |  |  |  |  |  |
| Autoimmune disease |  |  |  |  |  |  |  |  |  |  |  |  |
| 20 | nR | NS | Serious<sup>a</sup> | Serious<sup>b</sup> | Very<br>serious<sup>c</sup> | Strong<br>association | 1724/1979<br>(87.1) | 3943/4790<br>(82.3) | 1.07<br>(1.03,<br>1.11) | 6032.12<br>more per<br>100,000<br>(3068.95,<br>8995.3) | Type 4<sup>d</sup> | Critical |
| Solid organ transplant |  |  |  |  |  |  |  |  |  |  |  |  |
| 21 | nR | NS | Serious<sup>e</sup> | Serious<sup>b</sup> | Very<br>serious<sup>f</sup> | Strong<br>association | 1117/1852<br>(60.3) | 1442/2999<br>(48.1) | 1.29<br>(1.19,<br>1.39) | 14,229.94<br>more per<br>100,000<br>(11,269.98,<br>17,189.9) | Type 4<sup>d</sup> | Critical |
| Cancer (solid tumors and hematologic malignancies) |  |  |  |  |  |  |  |  |  |  |  |  |
| 30 | nR | NS | Serious<sup>g</sup> | Serious<sup>b</sup> | Very<br>serious<sup>h</sup> | Strong<br>association | 2362/3156<br>(74.8) | 2721/4232<br>(64.3) | 1.11<br>(1.06,<br>1.17) | 7860.6 more<br>per 100,000<br>(4543.3,<br>11,177.9) | Type 4<sup>d</sup> | Critical |
| Hematologic malignancy |  |  |  |  |  |  |  |  |  |  |  |  |
| 25 | nR | NS | Serious<sup>i</sup> | Serious<sup>b</sup> | NS | Strong<br>association | 2081/2803<br>(74.2) | 2271/3573<br>(63.6) | 1.16<br>(1.09,<br>1.24) | 10,490.65<br>more per<br>100,000<br>(6535.24,<br>14,446.05) | Type 3<sup>j</sup> | Critical |

GRADE, Grading of Recommendations, Assessment, Development, and Evaluation; nR, nonrandomized; NS, not serious; RoB, risk of bias; RR, risk ratio.
<sup>a</sup> $P=58.1\%$ , $X^2=45.38$ , $p(Q)=0$ ; moderate heterogeneity.
<sup>b</sup> Outcome definitions rather heterogeneous (various antibody types tested through many assays).
<sup>c</sup> Wider 95% CI due to only 3 events in the mRNA-1273 arm and overall small sample size in Hammer 2022.
<sup>d</sup>Lower grading due to imprecision and indirectness because of varying outcome definitions (different antibody types and assays used).
<sup>e</sup> $P=43.5\%$ , $X^2=35.42$ , $p(Q)=0.02$ ; no evidence of heterogeneity.
<sup>f</sup>Wider 95% CI due to only 1 event in the mRNA-1273 arm in Gallais 2022 and Maillard 2022 and the overall small sample size of the mRNA-1273 arm of Werbel 2021.
<sup>g</sup> $P=59.2\%$ , $X^2=71.11$ , $p(Q)=0$ ; moderate heterogeneity.
<sup>h</sup>Wider 95% CI due to only 2 events in the mRNA-1273 arm and overall small sample size of Macrae 2022.
<sup>i</sup> $P=57.0\%$ , $X^2=55.86$ , $p(Q)=0$ ; moderate heterogeneity.
<sup>j</sup>Higher grading due to strong association in RR. No issues with imprecision. Type 3 due to nonrandomized studies.

GRADE analysis found that the certainty of evidence of the seroconversion outcome overall and in the solid organ transplant and autoimmunity subgroups was downgraded to type 4 (very low) due to imprecision and indirectness because different assays and antibody thresholds were used to assess and define seroconversion across the studies analyzed. Evidence certainty in the subgroup of patients with hematologic malignancies was retained as type 3 (ie, the maximum certainty possible for nonrandomized studies) because there was a strong association in seroconversion likelihood and no issues with imprecision.

### Total Anti-spike Binding Antibody or IgG Titers

mRNA-1273 elicited a statistically significant relative increase in total anti-spike binding antibody or IgG titers versus BNT162b2 (50.45% [95% CI, 34.63%, 66.28%]; *P*<0.0001) in 45 nonrandomized studies of IC patients overall (**Table 1**). However, heterogeneity was considerable between studies (*I*^2^=89.5%).

Subgroup analyses showed that patients with solid organ transplant, solid tumors, and autoimmune disease vaccinated with mRNA-1273 compared with BNT162b2 had 335.04%, 104.82%, and 74.75% higher relative increases in total anti-spike binding antibody or IgG titers, respectively (**Table 3**). These findings were statistically significant (all *P*<0.05). Moderate and considerable heterogeneity was found in the solid organ transplant (*I*^2^=64.7%) and autoimmune disease (*I*^2^=91.8%) subgroups, with no evidence of heterogeneity in patients with solid tumors (*I*^2^=39.6%). Compared with BNT162b2, a higher relative increase in total anti-spike binding antibody or IgG titers in patients with hematologic malignancies was observed in the mRNA-1273 group; however this finding was not statistically significant (**Table 3**).

**Table 3.**
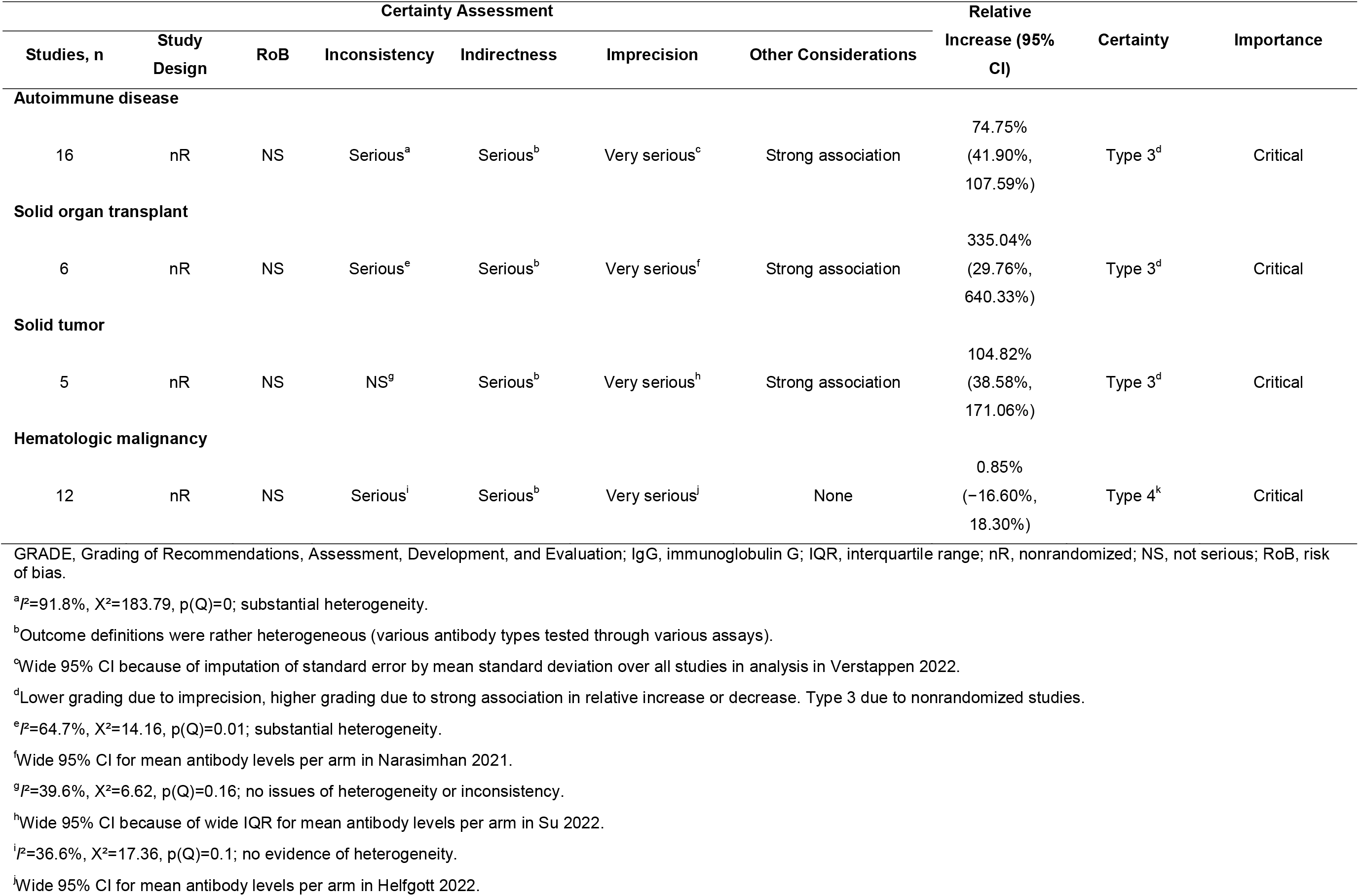
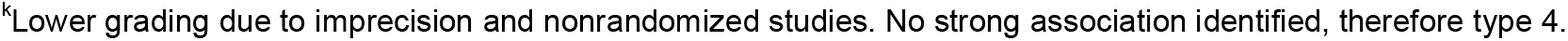
GRADE Summary of Total Anti-spike Binding Antibody and IgG Titers by Disease Subgroup.

GRADE evidence certainty for the total binding antibody or IgG titer outcome in IC patients overall and by autoimmune disease, solid organ transplant, and solid tumor subgroups was type 3 (low); lower grading because of imprecision was offset by higher grading due to a strong association in the relative increase in total anti-spike binding antibody or IgG titers.

### Neutralizing Anti-spike Titers

The SARS-CoV-2 variant against which the neutralization assays were performed was not reported in 5 studies [80, 82-85]; neutralization was evaluated against delta, omicron, and early WA1/2020 variants in 1 study [86] and against delta, omicron, and D614G/ancestral variants in the remaining study [87]. When assessed in IC patients overall from 7 nonrandomized studies, neutralizing anti-spike titers were not statistically significantly different in patients who received mRNA-1273 compared with BNT162b2 (**Table 1**). mRNA-1273 was associated with statistically significant relative increases in neutralizing anti-spike titers compared with BNT162b2 (436.78% [95% CI, 164.47%, 709.08%]; *P*=0.0017) in the subgroup of solid organ transplant recipients (**Table 4**). No evidence of heterogeneity was observed (*I*^2^=0%). No statistically significant differences in neutralizing anti-spike titers between vaccines were observed for the remaining subgroups (**Table 4**).

**Table 4.**
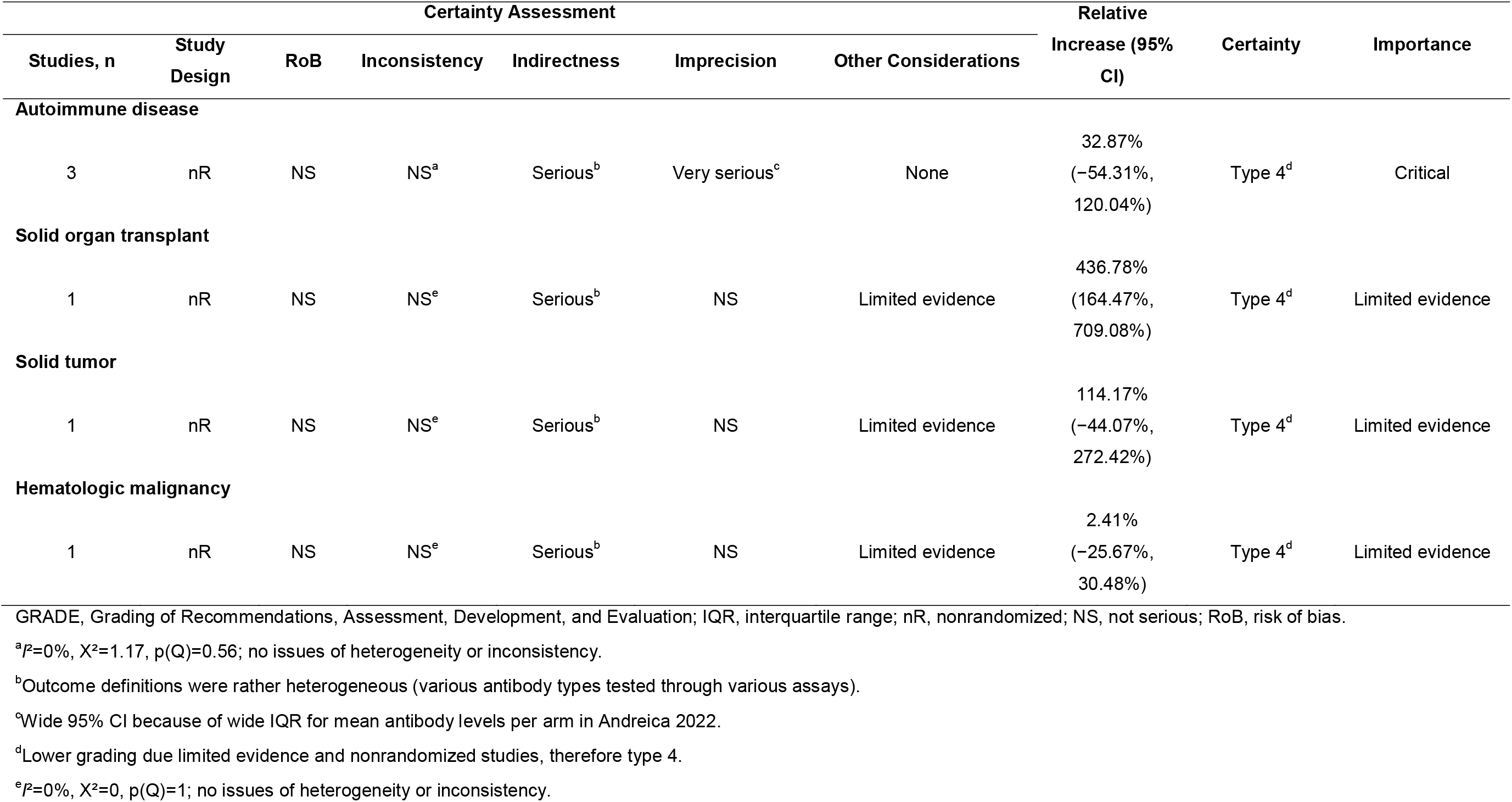
GRADE Summary of Neutralizing Anti-spike Titers by Disease Subgroup.

GRADE analysis found the certainty of evidence for the neutralizing anti-spike titer outcome in IC patients overall to be type 4 (very low) because of imprecision among the nonrandomized studies.

### Cellular Immune Response

The meta-analysis of 14 nonrandomized studies reporting cellular immune responses in IC patients overall found no statistically significant differences in the relative increase or decrease in the cellular immune response elicited by mRNA-1273 versus BNT162b2 (**Table 1**). Consistent with the meta-analysis of cellular immune response in IC patients overall, no statistically significant findings were observed in disease subgroups (**Table 5**).

**Table 5.**
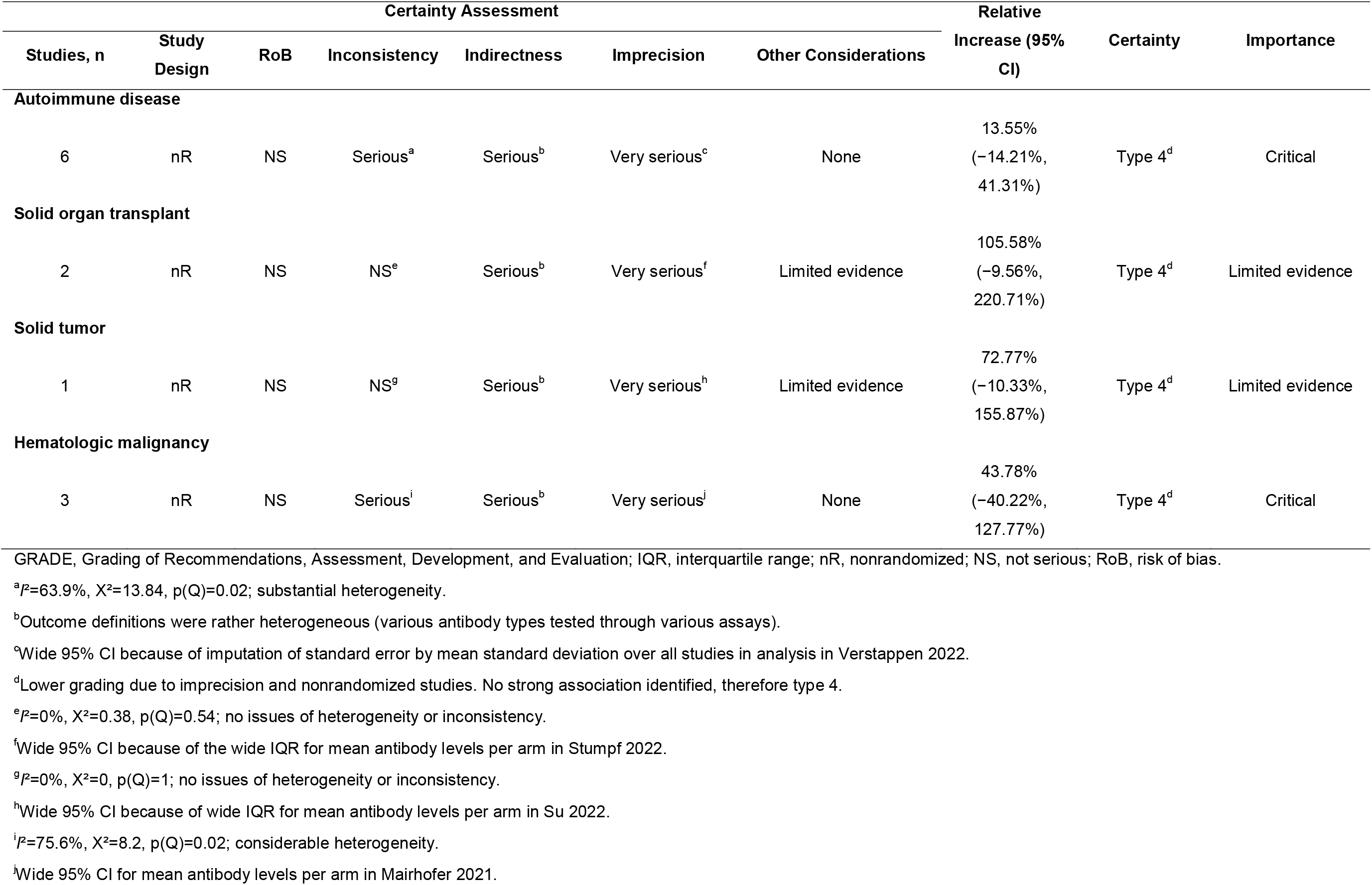
GRADE Summary of Findings for Cellular Immune Response by Disease Subgroup.

### Effect of Anti-CD20 Treatment on Outcomes

B-cell−depleting therapy through anti-CD20−specific monoclonal antibodies used to treat some patients with autoimmune disease or hematologic malignancies can severely impair the development of immune responses [88]. Therefore, and to account for heterogeneity between studies, we performed a subgroup analysis of patients with and without anti-CD20 monoclonal antibody treatment overall and in patients with autoimmune disease and hematologic malignancies. Vaccination with mRNA-1273 was associated with higher rates of seroconversion (**Table S9**) and elicited higher relative increases in total anti-spike binding antibody or IgG titers (**Table S10**), neutralizing anti-spike antibody titers (**Table S11**), and cellular immune responses (**Table S12**) in IC patients who received anti-CD20 treatment compared with BNT162b2. Generally similar trends were observed in patients with autoimmune disease and hematologic malignancies.

## Discussion

In this SLR and pairwise meta-analysis of IC patients ≥18 years of age vaccinated with mRNA COVID-19 vaccines, IC patients who received mRNA-1273 were significantly more likely to achieve seroconversion than those who received BNT162b2. Consistent with the seroconversion outcome, mRNA-1273 was also associated with a statistically significant relative increase in total anti-spike binding antibody or IgG titers over BNT162b2. Neutralizing anti-spike antibody titers were approximately 35% higher in IC patients vaccinated with mRNA-1273 versus BNT162b2; however, this result was not statistically significant. We also assessed cellular immune responses, which have been associated with protection against initial SARS-CoV-2 infection and viral clearance and may be critical for long-lasting immunity to SARS-CoV-2 [89]. Although no statistically significant differences in cellular immunity were observed between mRNA-1273 and BNT162b2 in this meta-analysis, the cellular immune response was approximately 15% higher in IC patients overall who were vaccinated with mRNA-1273 versus BNT162b2. Because only 14 nonrandomized studies, each with differing metrics of cellular immunity, were included in the meta-analysis for this outcome, it is possible that analyzing a larger number of studies with similar outcome definitions may have yielded clearer results.

Consistent with the practices of ACIP, the GRADE framework was used to evaluate evidence in this meta-analysis. Evidence for mRNA-1273 versus BNT162b2 vaccination was rated to be type 4 (very low) for the seroconversion, neutralizing antibody titer, and cellular immune response outcomes and type 3 (low) for the total antibody titer outcome (**Table 6**). Because IC populations are often excluded from RCTs, all but 1 of the 98 studies included in our meta-analysis were observational, nonrandomized studies, so the maximum evidence certainty achievable was type 3 (GRADE sets the maximum certainty for evidence derived from nonrandomized studies to type 3 [low] [47]). Logistical challenges, such as the rapid development and administration of mRNA COVID-19 vaccines in both general and IC populations, meant that RCTs designed specifically to test comparative immunogenicity of mRNA COVID-19 vaccines in IC patients were not feasible to conduct. Despite obligatory reliance on nonrandomized studies, we were able to consistently show strong associations between mRNA-1273 and the likelihood of achieving seroconversion and relative increases in total antibody titers in the observational studies identified in the SLR.

**Table 6.** Summary of Evidence for Outcomes of Interest.

| **Outcome** | **Outcome Importance<sup>a</sup>** | **Included in Evidence Profile** | **Certainty** |
| --- | --- | --- | --- |
| Seroconversion | Critical | Yes | Type 4 (very low) |
| Total anti-spike binding antibody or IgG titers | Critical | Yes | Type 3 (low) |
| Neutralizing anti-spike antibody titers | Critical | Yes | Type 4 (very low) |
| Cellular immune response | Critical | Yes | Type 4 (very low) |

GRADE, Grading of Recommendations, Assessment, Development, and Evaluation; IgG, immunoglobulin G.
<sup>a</sup>Relative importance of outcomes assessed and ranked per the GRADE framework.

Anti-CD20 monoclonal antibody therapy is used to treat some autoimmune diseases (eg, multiple sclerosis) [90] and hematologic malignancies characterized by aberrant B-cell proliferation (eg, non-Hodgkin lymphoma) [91]. Anti-CD20 therapy depletes B cells, possibly rendering patients receiving this therapy susceptible to SARS-CoV-2 infection and severe COVID-19 because of impaired humoral immune responses [88]. Our meta-analysis included studies of patients both with and without anti-CD20 treatment, which, while increasing the heterogeneity observed in some outcomes, allowed the impact of anti-CD20 treatment on differential immunogenicity of mRNA-1273 and BNT162b2 to be analyzed. Consistent with the overall IC population, mRNA-1273 was more likely to result in seroconversion and induce higher humoral and cellular immune responses than BNT162b2 in patients with anti-CD20 treatment. Although our findings were not statistically significant, in the absence of robust RCTs evaluating mRNA COVID-19 vaccines in IC populations receiving anti-CD20 therapy, they suggest that mRNA-1273 may be more immunogenic than BNT162b2 in this population at even greater risk for severe COVID-19 compared with the overall IC population.

Limitations of our study are that non-English studies were excluded from the SLR and that publication bias was not assessed in the meta-analysis. Furthermore, we observed considerable heterogeneity, in part due to differing study designs (eg, case control, cohort), differences in the number of vaccine doses or booster doses administered by study as well as lack of detail regarding mRNA dosage in reported mRNA-1273 booster doses, differing outcome definitions between studies, and differences in methods and assays used to measure antibody titers. Changing prevalence of variants of concern over time and differing immune responses to the variants as well as the heterogenous nature of IC conditions and background treatments (eg, anti-CD20 therapy, immunoglobulin replacement therapy) also contributed to the heterogeneity we observed. To address some of these limitations, we conducted subgroup analyses by IC condition and anti-CD20 treatment subgroups as previously discussed, preferentially included antibody titers from the Roche Elecsys platform where available, and used relative increase or decrease in humoral and cellular immunity outcomes to mitigate the heterogeneity caused by multiple outcome definitions and cutoffs. Although these strategies reduced heterogeneity, more data are needed to address all the limitations of our meta-analysis.

As previously described [52], differences in prescribing behavior or the impact of vaccine choice could not be accounted for in our meta-analysis. Besides mRNA dose in mRNA-1273 and BNT162b2, other differences in vaccine formulation or delivery, such as the type of lipid nanoparticle encapsulating the mRNA, mRNA translation efficiency, and vaccination schedule (4 vs 3 weeks between doses of the primary series), may have impacted immunogenicity.

In conclusion, our meta-analysis of mostly observational studies showed that vaccination with a higher-dose mRNA-1273 primary series (100 mcg mRNA/dose) or primary series and booster (50 or 100 mcg mRNA/dose) was more likely to result in seroconversion and elicited higher total anti-spike antibody titers than the BNT162b2 primary series or primary series and booster (30 mcg mRNA/dose irrespective of dose type). These findings suggest that immune responses to SARS-CoV-2 vaccination could be optimized in IC patients by selecting an appropriate mRNA vaccine and can help inform vaccine strategy in high-risk, immunosuppressed populations.

## Data Availability

All data produced in the present work are contained in the manuscript.

## Author Contributions

SK, KH, and XW designed and performed the systematic literature review and meta-analysis, interpreted data, and critically evaluated the manuscript. AS and AK designed and performed the systematic literature review and critically evaluated the manuscript. PS, MA, SV, MN, and NK collected data and critically evaluated the manuscript. MTB-J and NVdV conceptualized the article and provided oversight and critical evaluation of the manuscript. All authors contributed to the article and approved the submitted version.

## Acknowledgments

The authors thank Ekkehard Beck of Moderna, Inc., for critical review of the manuscript. Writing assistance was provided by Erin McClure, PhD, an employee of ICON (Blue Bell, PA, USA) in accordance with Good Publication Practice (GPP3) guidelines, funded by Moderna, Inc., and under the direction of the authors.

