## Supplemental Table 1 for "Immunogenicity of mRNA-1273 and BNT162b2 in Immunocompromised Patients: Systematic Review and Meta-Analysis Using GRADE"

### Table S1. Databases and Strategies Used for the Systematic Literature Review

| **Search No.** | **Query Scope** | **Query Script** |
| --- | --- | --- |
| *● Database:* WHO COVID-19 global literature on coronavirus disease including MEDLINE (PubMed)  *● URL:* <https://search.bvsalud.org/global-literature-on-novel-coronavirus-2019-ncov/> | | |
| 1 | All COVID-19 | Not applicable |
| 2 | Major focus on COVID-19 vaccine(s) or vaccination | mj: Covid-19 vacc* |
| 3 | Vaccine efficacy, effectiveness, related research types and terms | ("vaccine effectiveness" OR "vaccine efficacy" OR "vaccine research" OR "vaccine study" OR "vaccine trial" OR "vaccine control" ~2 OR "vaccine comparison"~2 OR "vaccine response" OR "IgM" OR "IgG" OR Ig* OR antibod* OR "anti-body" OR immunoglob* OR "immuno-globulin" OR immunogen* OR "immuno-genicity" OR sero* OR "immune response"~2) |
| 4 | At-risk medical conditions | ("risk condition"~2 OR "risk disorder"~2 OR "at-risk" ~2 OR "at risk" ~2 OR " high risk" OR "high-risk" OR "risk disease" ~2 OR "risk comorbidity" OR "risk disability" OR "long term condition"~2 OR "long-term condition"~2 OR underlying* OR pre-existing* OR preexisting* OR "medical condition" OR comorbid* OR immunocompr* OR immunodef* OR immunosupp* OR "immuno-compromised" OR "immuno-deficiency" OR "immuno-suppression" OR cancer* OR carcinoma* OR malignan* OR neoplasm* OR "solid organ" OR chemotherap* OR antineoplastic OR "anti-neoplastic" OR "cytotoxic therapy"~2 OR "anti-cancer" OR anticancer OR transplant* OR "stem cell" OR "SCT" OR "HSCT" OR "BMT" OR "chronic kidney" OR "CKD" OR "chronic liver" OR "CLD" OR cirrho* OR "chronic hepatitis" ~2 OR "HCC" OR haemochromatosis OR hemochromatosis OR "metabolic liver" OR "genetic liver" ~2 OR "chronic nephrotic"~2 OR "chronic hepatic"~2 OR asthma* OR "bronchiectasis" OR "bronchopulmonary dysplasia" OR "chronic lung" OR "chronic pulmonary" OR "chronic obstructive" OR "COPD" OR emphysema OR "chronic bronchitis" OR "interstitial lung disease" OR "idiopathic pulmonary fibrosis" OR "pulmonary embolism" OR "pulmonary hypertension" OR "cystic fibrosis" OR diabet* OR "IDDM" OR "NIDDM" OR "overweight" OR obes* OR dementia OR Alzheimer* OR "degenerative brain"~2 OR "degenerative mental"~2 OR neurolog* OR neurodegen* OR schizo* OR "psychosis" OR "psychotic" OR "severe mood disorder" OR "clinical depression" OR "substance use" OR "substance abuse" OR "attention-deficit/hyperactivity disorder" OR "attention disorder"~2 OR "ADHD" OR "spinal cord injury" OR "cerebral palsy" OR "birth defect" ~2 OR "birth anomaly" OR "intellectual and developmental disability" OR "IDD" OR "learning disability" OR "Down syndrome" OR "inborn error" OR congenital OR inborn OR "inherited abnormality" OR "inherited disorder" OR "genetic abnormality"~2 OR "genetic disorder" ~2 OR heart OR cardiac OR coronary OR cardio* OR "CHD" OR "high blood pressure" OR "hypertension" OR myocard* OR cardiovasc* OR "HIV" OR "AIDS" OR "inherited red blood cell disorders" ~3 OR haemoglobinopath* OR hemoglobinopath* OR hemolytic OR haemolytic OR hematologic* OR haematologic* OR "stroke" OR "cerebrovascular" OR "CVD" OR "tuberculosis" OR "TB") |
| 5 | Without filtering for vaccine efficacy etc (#2 AND #4) Major focus on COVID-19 vaccine(s) or vaccination  AND At-risk medical conditions | (mj: Covid-19 vacc*) AND ("risk condition"~2 OR "risk disorder"~2 OR "at-risk" ~2 OR "at risk" ~2 OR " high risk" OR "high-risk" OR "risk disease" ~2 OR "risk comorbidity" OR "risk disability" OR "long term condition"~2 OR "long-term condition"~2 OR underlying* OR pre-existing* OR preexisting* OR "medical condition" OR comorbid* OR immunocompr* OR immunodef* OR immunosupp* OR "immuno-compromised" OR "immuno-deficiency" OR "immuno-suppression" OR cancer* OR carcinoma* OR malignan* OR neoplasm* OR "solid organ" OR chemotherap* OR antineoplastic OR "anti-neoplastic" OR "cytotoxic therapy"~2 OR "anti-cancer" OR anticancer OR transplant* OR "stem cell" OR "SCT" OR "HSCT" OR "BMT" OR "chronic kidney" OR "CKD" OR "chronic liver" OR "CLD" OR cirrho* OR "chronic hepatitis" ~2 OR "HCC" OR haemochromatosis OR hemochromatosis OR "metabolic liver" OR "genetic liver" ~2 OR "chronic nephrotic"~2 OR "chronic hepatic"~2 OR asthma* OR "bronchiectasis" OR "bronchopulmonary dysplasia" OR "chronic lung" OR "chronic pulmonary" OR "chronic obstructive" OR "COPD" OR emphysema OR "chronic bronchitis" OR "interstitial lung disease" OR "idiopathic pulmonary fibrosis" OR "pulmonary embolism" OR "pulmonary hypertension" OR "cystic fibrosis" OR diabet* OR "IDDM" OR "NIDDM" OR "overweight" OR obes* OR dementia OR Alzheimer* OR "degenerative brain"~2 OR "degenerative mental"~2 OR neurolog* OR neurodegen* OR schizo* OR "psychosis" OR "psychotic" OR "severe mood disorder" OR "clinical depression" OR "substance use" OR "substance abuse" OR "attention-deficit/hyperactivity disorder" OR "attention disorder"~2 OR "ADHD" OR "spinal cord injury" OR "cerebral palsy" OR "birth defect" ~2 OR "birth anomaly" OR "intellectual and developmental disability" OR "IDD" OR "learning disability" OR "Down syndrome" OR "inborn error" OR congenital OR inborn OR "inherited abnormality" OR "inherited disorder" OR "genetic abnormality"~2 OR "genetic disorder" ~2 OR heart OR cardiac OR coronary OR cardio* OR "CHD" OR "high blood pressure" OR "hypertension" OR myocard* OR cardiovasc* OR "HIV" OR "AIDS" OR "inherited red blood cell disorders" ~3 OR haemoglobinopath* OR hemoglobinopath* OR hemolytic OR haemolytic OR hematologic* OR haematologic* OR "stroke" OR "cerebrovascular" OR "CVD" OR "tuberculosis" OR "TB") |
| 6 | Full query (#2 AND #3 AND #4) Major focus on COVID-19 vaccine(s) or vaccination AND Vaccine efficacy, effectiveness, related research types and terms AND At-risk medical conditions | (mj: Covid-19 vacc*) AND ("vaccine effectiveness" OR "vaccine efficacy" OR "vaccine research" OR "vaccine study" OR "vaccine trial" OR "vaccine control" ~2 OR "vaccine comparison"~2 OR "vaccine response" OR "IgM" OR "IgG" OR Ig* OR antibod* OR "anti-body" OR immunoglob* OR "immuno-globulin" OR immunogen* OR "immuno-genicity" OR sero* OR "immune response"~2) AND ("risk condition"~2 OR "risk disorder"~2 OR "at-risk" ~2 OR "at risk" ~2 OR " high risk" OR "high-risk" OR "risk disease" ~2 OR "risk comorbidity" OR "risk disability" OR "long term condition"~2 OR "long-term condition"~2 OR underlying* OR pre-existing* OR preexisting* OR "medical condition" OR comorbid* OR immunocompr* OR immunodef* OR immunosupp* OR "immuno-compromised" OR "immuno-deficiency" OR "immuno-suppression" OR cancer* OR carcinoma* OR malignan* OR neoplasm* OR "solid organ" OR chemotherap* OR antineoplastic OR "anti-neoplastic" OR "cytotoxic therapy"~2 OR "anti-cancer" OR anticancer OR transplant* OR "stem cell" OR "SCT" OR "HSCT" OR "BMT" OR "chronic kidney" OR "CKD" OR "chronic liver" OR "CLD" OR cirrho* OR "chronic hepatitis" ~2 OR "HCC" OR haemochromatosis OR hemochromatosis OR "metabolic liver" OR "genetic liver" ~2 OR "chronic nephrotic"~2 OR "chronic hepatic"~2 OR asthma* OR "bronchiectasis" OR "bronchopulmonary dysplasia" OR "chronic lung" OR "chronic pulmonary" OR "chronic obstructive" OR "COPD" OR emphysema OR "chronic bronchitis" OR "interstitial lung disease" OR "idiopathic pulmonary fibrosis" OR "pulmonary embolism" OR "pulmonary hypertension" OR "cystic fibrosis" OR diabet* OR "IDDM" OR "NIDDM" OR "overweight" OR obes* OR dementia OR Alzheimer* OR "degenerative brain"~2 OR "degenerative mental"~2 OR neurolog* OR neurodegen* OR schizo* OR "psychosis" OR "psychotic" OR "severe mood disorder" OR "clinical depression" OR "substance use" OR "substance abuse" OR "attention-deficit/hyperactivity disorder" OR "attention disorder"~2 OR "ADHD" OR "spinal cord injury" OR "cerebral palsy" OR "birth defect" ~2 OR "birth anomaly" OR "intellectual and developmental disability" OR "IDD" OR "learning disability" OR "Down syndrome" OR "inborn error" OR congenital OR inborn OR "inherited abnormality" OR "inherited disorder" OR "genetic abnormality"~2 OR "genetic disorder" ~2 OR heart OR cardiac OR coronary OR cardio* OR "CHD" OR "high blood pressure" OR "hypertension" OR myocard* OR cardiovasc* OR "HIV" OR "AIDS" OR "inherited red blood cell disorders" ~3 OR haemoglobinopath* OR hemoglobinopath* OR hemolytic OR haemolytic OR hematologic* OR haematologic* OR "stroke" OR "cerebrovascular" OR "CVD" OR "tuberculosis" OR "TB") |
| 7 | Full query (#2 AND #3 AND #4) Major focus on COVID-19 vaccine(s) or vaccination AND Vaccine efficacy, effectiveness, related research types and terms AND At-risk medical conditions  English publications | (mj: covid-19 vacc*) AND ("vaccine effectiveness" OR "vaccine efficacy" OR "vaccine research" OR "vaccine study" OR "vaccine trial" OR "vaccine control" ~2 OR "vaccine comparison"~2 OR "vaccine response" OR "IgM" OR "IgG" OR ig* OR antibod* OR "anti-body" OR immunoglob* OR "immuno-globulin" OR immunogen* OR "immuno-genicity" OR sero* OR "immune response"~2) AND ("risk condition"~2 OR "risk disorder"~2 OR "at-risk" ~2 OR "at risk" ~2 OR " high risk" OR "high-risk" OR "risk disease" ~2 OR "risk comorbidity" OR "risk disability" OR "long term condition"~2 OR "long-term condition"~2 OR underlying* OR pre-existing* OR preexisting* OR "medical condition" OR comorbid* OR immunocompr* OR immunodef* OR immunosupp* OR "immuno-compromised" OR "immuno-deficiency" OR "immuno-suppression" OR cancer* OR carcinoma* OR malignan* OR neoplasm* OR "solid organ" OR chemotherap* OR antineoplastic OR "anti-neoplastic" OR "cytotoxic therapy"~2 OR "anti-cancer" OR anticancer OR transplant* OR "stem cell" OR "SCT" OR "HSCT" OR "BMT" OR "chronic kidney" OR "CKD" OR "chronic liver" OR "CLD" OR cirrho* OR "chronic hepatitis" ~2 OR "HCC" OR haemochromatosis OR hemochromatosis OR "metabolic liver" OR "genetic liver" ~2 OR "chronic nephrotic"~2 OR "chronic hepatic"~2 OR asthma* OR "bronchiectasis" OR "bronchopulmonary dysplasia" OR "chronic lung" OR "chronic pulmonary" OR "chronic obstructive" OR "COPD" OR emphysema OR "chronic bronchitis" OR "interstitial lung disease" OR "idiopathic pulmonary fibrosis" OR "pulmonary embolism" OR "pulmonary hypertension" OR "cystic fibrosis" OR diabet* OR "IDDM" OR "NIDDM" OR "overweight" OR obes* OR dementia OR alzheimer* OR "degenerative brain"~2 OR "degenerative mental"~2 OR neurolog* OR neurodegen* OR schizo* OR "psychosis" OR "psychotic" OR "severe mood disorder" OR "clinical depression" OR "substance use" OR "substance abuse" OR "attention-deficit/hyperactivity disorder" OR "attention disorder"~2 OR "ADHD" OR "spinal cord injury" OR "cerebral palsy" OR "birth defect" ~2 OR "birth anomaly" OR "intellectual and developmental disability" OR "IDD" OR "learning disability" OR "Down syndrome" OR "inborn error" OR congenital OR inborn OR "inherited abnormality" OR "inherited disorder" OR "genetic abnormality"~2 OR "genetic disorder" ~2 OR heart OR cardiac OR coronary OR cardio* OR "CHD" OR "high blood pressure" OR "hypertension" OR myocard* OR cardiovasc* OR "HIV" OR "AIDS" OR "inherited red blood cell disorders" ~3 OR haemoglobinopath* OR hemoglobinopath* OR hemolytic OR haemolytic OR hematologic* OR haematologic* OR "stroke" OR "cerebrovascular" OR "CVD" OR "tuberculosis" OR "TB") AND la:("en") |
| *● Database:* WHO COVID-19 global literature on coronavirus disease excluding MEDLINE (Pubmed), including other reference sources in the WHO COVID-19 database (ICTRP, EMBASE, EuropePMC, PREPRINT-MEDRXIV, Web of Science, ProQuest Central, Academic Search Complete, Scopus, COVIDWHO) – full query #3  *● URL:* <https://search.bvsalud.org/global-literature-on-novel-coronavirus-2019-ncov/> | | |
| 1 | All COVID-19 | Not applicable |
| 2 | WHO-accessed databases other than MEDLINE (PubMed) | db:("ICTRP" OR "EMBASE" OR "EuropePMC" OR "PREPRINT-MEDRXIV" OR "Web of Science" OR "ProQuest Central" OR "Academic Search Complete" OR "Scopus" OR "COVIDWHO") |
| 3 | COVID-19 vaccine(s) or vaccination AND vaccine efficacy, effectiveness, related research types and terms AND  At-risk medical conditions AND WHO accessed databases other than MEDLINE (PubMed) | ("covid19 vaccine" ~2 OR "covid-19 vaccine"~2 OR "covid-19 vaccines"~2 OR "covid19 vaccines" ~2 OR "covid-19 vaccination"~2 OR "Covid19 vaccination"~2) AND ("vaccine effectiveness" OR "vaccine efficacy" OR "vaccine research" OR "vaccine study" OR "vaccine trial" OR "vaccine control" ~2 OR "vaccine comparison"~2 OR "vaccine response" OR "IgM" OR "IgG" OR Ig* OR antibod* OR "anti-body" OR immunoglob* OR "immuno-globulin" OR immunogen* OR "immuno-genicity" OR sero* OR "immune response"~2) AND ("risk condition"~2 OR "risk disorder"~2 OR "risk disease" ~2 OR "risk comorbidity" OR "risk disability" or "long term condition"~2 OR "long-term condition"~2 OR underlying* OR pre-existing* OR preexisting* OR "medical condition" OR comorbid* OR immunocompr* OR immunodef* or immunosupp* OR "immuno-compromised" OR "immuno-deficiency" OR "immuno-suppression" or cancer* OR carcinoma* OR malignan* OR neoplasm* OR "solid organ" OR chemotherap* OR antineoplastic OR "anti-neoplastic" OR "cytotoxic therapy"~2 OR "anti-cancer" OR anticancer or transplant* OR "stem cell" OR "SCT" OR "HSCT" OR "BMT" OR "chronic kidney" OR "CKD" OR "chronic liver" OR "CLD" OR cirrho* OR "chronic hepatitis" ~2 OR "HCC" OR haemochromatosis OR hemochromatosis or "metabolic liver" OR "genetic liver" ~2 OR "chronic nephrotic"~2 OR "chronic hepatic"~2 OR asthma* OR "bronchiectasis" OR "bronchopulmonary dysplasia" OR "chronic lung" OR "chronic pulmonary" or "chronic obstructive" OR "COPD" OR emphysema OR "chronic bronchitis" OR "interstitial lung disease" OR "idiopathic pulmonary fibrosis" OR "pulmonary embolism" OR "pulmonary hypertension" OR "cystic fibrosis" OR diabet* OR "IDDM" OR "NIDDM" OR "overweight" OR obes* OR dementia OR alzheimer* OR "degenerative mental"~2 OR neurolog* OR neurodegen* OR schizo* or "psychosis" OR "psychotic" OR "severe mood disorder" OR "clinical depression" OR "substance use" OR "substance abuse" OR "attention-deficit/hyperactivity disorder" OR "attention disorder"~2 or "ADHD" OR "spinal cord injury" OR "cerebral palsy" OR "birth defect" ~2 OR "birth anomaly" OR "intellectual and developmental disability" OR "IDD" OR "learning disability" OR "Down syndrome" or "inborn error" OR congenital OR inborn OR "inherited abnormality" OR "inherited disorder" OR "genetic abnormality"~2 OR "genetic disorder" ~2 OR heart OR cardiac OR coronary OR cardio* OR "CHD" or "high blood pressure" OR "hypertension" OR myocard* OR cardiovasc* OR "HIV" OR "AIDS" OR "inherited red blood cell disorders" ~3 OR haemoglobinopath* OR hemoglobinopath* OR hemolytic OR haemolytic OR hematologic* OR haematologic* OR "stroke" OR "cerebrovascular" OR "CVD" OR "tuberculosis" OR "TB") AND db:("ICTRP" OR "EMBASE" OR "EuropePMC" OR "PREPRINT-MEDRXIV" OR "Web of Science" OR "ProQuest Central" OR "Academic Search Complete" OR "Scopus" OR "COVIDWHO") |
