## Supplemental Table 2 for "Immunogenicity of mRNA-1273 and BNT162b2 in Immunocompromised Patients: Systematic Review and Meta-Analysis Using GRADE"

### Table S2. Research Question and PICOS

| *● Research question:* Does the 2-dose mRNA-1273 COVID-19 vaccine primary series (100 mcg mRNA/dose) or primary series and booster (50 or 100 mcg mRNA/dose) have greater immunogenicity in IC populations compared with the 2-dose BNT162b2 COVID-19 vaccine primary series or primary series and booster (30 mcg mRNA/dose irrespective of dose type)? | | |
| --- | --- | --- |
| **PICOS** | **Includes** | **Excludes** |
| Population | - IC individuals ≥18 years of age defined as people with medical conditions associated with CEV groups 1 and 2^a^ [9] | - Studies in pregnant women, current/former smokers, physically inactive individuals - Studies in only healthy individuals or individuals not categorized as CEV |
| Intervention/Exposure | - mRNA-1273 | - Studies with heterologous vaccination schedule (i.e., data on mixed mRNA-1273, BNT162b2, or other vaccines) |
| Comparison | - BNT162b2 |  |
| Outcomes | - Seroconversion - Total anti-spike binding antibody or IgG titers - Neutralizing anti-spike antibody titers - Cellular immune response | - Studies with only safety results |
| Study design | - Clinical trials - Observational studies - Any kind of real-world evidence | - Study protocol (no results) - Economic models |
| Other limits | - Any publication type (including letters, commentary, abstract, full text, poster) - Publication in English language | - Non–English language publications |

CEV, clinically extremely vulnerable; COVID-19, coronavirus disease 2019; IC, immunocompromised; IgG, immunoglobulin; PICOS, population, intervention, comparison, outcomes, and study design.

^a^CEV groups 1 or 2 comprise transplant recipients, patients with cancer, primary immunodeficiencies, dialysis or severe kidney disease, poorly controlled HIV infection, or autoimmune diseases requiring immunosuppressive therapy.
