## Supplemental Table 3 for "Immunogenicity of mRNA-1273 and BNT162b2 in Immunocompromised Patients: Systematic Review and Meta-Analysis Using GRADE"

### Table S3. Characteristics of Studies Included in the Meta-Analysis: Seroconversion

| **Author, Year** | **Study Characteristics** | | | | | | |  | **Vaccinated, n** | |  | **Seroconversion, n/N (%)** | | **RoB** |
| --- | --- | --- | --- | --- | --- | --- | --- | --- | --- | --- | --- | --- | --- | --- |
|  | **Study Design** | **Population** | **Anti-CD20 mAb** | **Vaccination** | **Study Period** | **Assay/ Platform** | **Seropositivity Definition** |  | **mRNA-1273** | **BNT 162b2** |  | **mRNA-1273** | **BNT 162b2** |  |
| Abid, 2022 [66] | Retrospective cohort | - USA - Recipients of HSCT, CAR T cell, or BiTE | No | MMM vs PPP | NR | Abbott AdviseDx SARS-CoV-2 IgG II | Anti-S1 RBD IgG titers ≥50 AU/mL |  | 23 | 43 |  | 13/23 (56.5) | 27/43 (62.8) | S |
| Addeo, 2021 [92] | Prospective observational cohort | - Switzerland, USA - Patients with cancer | Yes | MM vs PP | Jan–Apr 2021 | Roche Elecsys | Anti-spike RBD titers ≥0.8 U/mL |  | 93 | 30 |  | 88/93 (94.6) | 28/30 (93.3) | NS |
| Agha, 2021 [93] | Retrospective cross-sectional | - USA - Patients with hematologic malignancies | Yes | MM vs PP | Dec 2020–Mar 2021 | Beckman Coulter SARS-CoV-2 | Anti-spike RBD IgG extinction coefficient ≥1.00 |  | 28 | 34 |  | 16/28 (57.1) | 15/34 (44.1) | S |
| Aleman, 2021 [94] | Retrospective cohort | - USA - Patients with multiple myeloma | No | MM vs PP | NR | NR | Anti-spike IgG >5 AU/mL |  | 37 | 99 |  | 33/37 (89.2) | 81/99 (81.8) | S |
| Anand, 2021 [95] | Prospective cohort | - USA - Patients receiving dialysis | No | MM vs PP | Aug 2020–Apr 2021 | Siemens semi-quantitative | Anti-RBD IgG index value ≥10 |  | 353 | 239 |  | 321/353 (90.9) | 185/239 (77.4) | NS |
| Arranz, 2022 [96] | Prospective observational cohort | - Spain - Patients with IBD | No | MM vs PP | 15 Feb–30 Jul 2021 | Siemens Healthineers and Vircell | Anti-spike IgG cutoff >1, Anti-N IgG cutoff >1.6 |  | 18 | 144 |  | 17/18 (94.4) | 134/144 (93.1) | S |
| Arroyo-Sánchez, 2022 [97] | Prospective cohort | - Spain - Patients with common variable immunodeficiency | Yes | MM vs PP | Feb-Jun 2021 | EUROIMMUN ELISA | Anti-S1 IgG relative OD ≥1.1 |  | 5 | 12 |  | 5/5 (100.0) | 9/12 (75.0) | NS |
| Arumahandi de Silva, 2022 [98] | Retrospective cohort | - Germany - Patients with autoimmune rheumatic diseases receiving methotrexate | No | MM vs PP | Apr–Sep 2021 | SeraSpot anti-SARS-CoV-2 IgG microarray-based immunoassay | Anti-RBD IgG signal-to-cutoff ratio >1.00 |  | 8 | 47 |  | 7/8 (87.5) | 35/47 (74.5) | NS |
| Bae, 2022 [99] | Prospective cohort | - USA - Solid organ transplant recipients | No | MM vs PP | Dec 2020–May 2021 | Roche Elecsys, EUROIMMUN ELISA | Anti-spike RBD titers ≥0.8 U/mL; Anti-S1 IgG titers ≤1.1 AU/mL |  | 478 | 559 |  | 311/478 (65.1) | 293/559 (52.4) | NS |
| Bagacean, 2022 [67] | Retrospective cohort | - France - Patients with CLL | Yes | MMM vs PPP | Mar–Jul 2021 | Abbott AdviseDx SARS-CoV-2 IgG II | Anti-spike IgG  titers ≥50 AU/mL |  | 69 | 367 |  | 47/69 (68.1) | 187/367 (51.0) | S |
| Barmettler, 2022 [68] | Retrospective longitudinal cohort | - USA - Patients with predominant antibody deficiency | Yes | MMM vs PPP | Dec 2020–Jun 2021 | Roche Elecsys | Anti-spike RBD titers ≥100 U/mL |  | 33 | 25 |  | 23/33 (69.7) | 14/25 (56.0) | S |
| Branagan, 2021 [100] | Prospective clinical trial | - USA - Patients with multiple myeloma and Waldenström macroglobulinemia | No | MM vs PP | NR | Roche Elecsys | Anti-spike RBD  titers ≥100 U/mL |  | 34 | 47 |  | 23/34 (67.6) | 18/47 (38.3) | N/A |
| Broseta, 2021 [101] | Prospective observational cohort | - Spain - Patients receiving hemodialysis | No | MM vs PP | Feb–Apr 2021 | Siemens Healthineers Atellica IM | Anti-S1 RBD IgG titers ≥1 U/mL |  | 100 | 75 |  | 98/100 (98.0) | 69/75 (92.0) | NS |
| Chaekal, 2022 [69] | Retrospective cohort | - USA - Allogeneic transplant (using T-cell depletion) recipients | Yes | MMM vs PPP | Jan 2021–Jan 2022 | Roche Elecsys | Anti-spike RBD  titers >0.8 BAU/mL; >100 BAU/mL defined as “high responder” |  | 48 | 89 |  | 36/48 (75.0) | 57/89 (64.0) | NS |
| Chang, 2022 [86] | Observational cohort | - USA - Patients with NHL including CLL | Yes | MM vs PP | NR | NR | Anti-full length spike IgG titers ≥500 AU/mL |  | 53 | 54 |  | 33/53 (62.3) | 39/54 (72.2) | NS |
| Chen, 2022 [102] | Prospective observational | - Taiwan - Kidney transplant recipients | No | MM vs PP | Jul–Dec 2021 | Roche Elecsys | Anti-spike RBD titers ≥0.8 U/mL |  | 76 | 34 |  | 58/76 (76.3) | 18/34 (52.9) | NS |
| Chiang, 2022 [70] | Prospective cohort | - USA - Solid organ transplant recipients | No | MMM vs PPP | Mar 2021–Jan 2022 | Roche Elecsys, EUROIMMUN ELISA | Anti-spike RBD titers ≥0.8 U/mL; Anti-S1 IgG ≤1.1 AU |  | 84 | 149 |  | 43/84 (51.2) | 78/149 (52.3) | NS |
| Connolly, 2022 [103] | Case series | - USA - Patients with rheumatic and musculoskeletal diseases who withheld mycophenolate | Yes | MM vs PP | 17 Dec 2020–13 May 2021 | Roche Elecsys | Anti-spike RBD  titers >0.79 U/mL |  | 8 | 13 |  | 7/8 (87.5) | 12/13 (92.3) | N/A |
| Debska-Slizien, 2022 [104] | Longitudinal | - Poland - Kidney transplant Recipients | No | MM vs PP | NR | DiaSorin LIAISON | Anti-trimeric S-domain IgG cutoff index value >12 AU/mL |  | 37 | 105 |  | 23/37 (62.2) | 49/105 (46.7) | NS |
| Doukas, 2022 [105] | Retrospective observational | - USA - Patients with CLL | Yes | MM vs PP | Jan–Oct 2021 | NR | Detectable anti-spike IgG and total antibody titers |  | 72 | 106 |  | 44/72 (61.1) | 47/106 (44.3) | S |
| Ferri, 2021 [106] | Prospective cohort | - Italy - Patients with autoimmune systemic diseases | Yes | MM vs PP | NR | Abbott AdviseDx SARS-CoV-2 IgG II | Anti-trimeric S-domain IgG titers ≥7 BAU/mL |  | 30 | 448 |  | 27/30 (90.0) | 388/448 (86.6) | S |
| Floyd, 2022 [107] | Multicenter cohort | - USA, UK, Germany - Patients with antineutrophil cytoplasmic autoantibody–associated vasculitis | Yes | MM vs PP | Mar–Sep 2021 | Roche Elecsys, DiaSorin LIAISON, EUROIMMUN ELISA | Anti-spike RBD titers >1.0 U/mL (Roche; USA), >0.8 U/mL (DiaSorin), >1.24 U/mL (EUROIMMUN), >0.8 U/mL (Roche; UK), ≥0.8 U/mL (Roche; Germany) |  | 31 | 89 |  | 19/31 (61.3) | 49/89 (55.1) | NS |
| Fujieda, 2022 [108] | Retrospective cohort | - Japan - Kidney transplant recipients | No | MM vs PP | Jun–Sep 2021 | Abbott SARS-CoV-2 IgG II Quant | Anti-trimeric S-domain IgG titers ≥50 AU/mL |  | 13 | 52 |  | 8/13 (61.5) | 14/52 (26.9) | S |
| Gallais, 2022 [79] | Retrospective cohort | - France - COVID-19–naive lung transplant recipients | No | MMM vs PPP | Apr–Oct 2021 (2–8 weeks after third dose) | Abbott AdviseDx SARS-CoV-2 IgG II | Anti-spike RBD IgG  titers ≥7.1 BAU/mL |  | 41 | 13 |  | 6/41 (14.6) | 1/13 (7.7) | NS |
| Giannella, 2022 [109] | Prospective observational multicenter cohort | - Italy and Spain - Solid organ transplant recipients | No | MM vs PP | Jan–Nov 2021 | Roche Elecsys, MSD | Anti-spike RBD titers ≥5 U/mL (Roche) or ≥45 BAU/mL (MSD) |  | 134 | 928 |  | 93/134 (69.4) | 463/928 (49.9) | NS |
| Göschl, 2022 [110] | Retrospective cohort | - Austria - Patients with inborn errors of immunity | Yes | MM vs PP | NR | Roche Elecsys (qualitative) | Anti-spike RBD titers >1,000 cutoff index |  | 6 | 17 |  | 4/6 (66.7) | 13/17 (76.5) | NS |
| Greenberger, 2021 [111] | Prospective cohort registry | - USA - Patients with hematologic malignancies | Yes | MM vs PP | Mar–May 2021 | Roche Elecsys | Anti-spike RBD  titers ≥0.8 U/mL |  | 652 | 793 |  | 505/652 (77.5) | 583/793 (73.5) | S |
| Greenberger, 2022 [112] | Prospective cohort registry | - USA - Patients with hematologic malignancies | Yes | MM vs PP | May–Dec 2021 | Roche Elecsys | Anti-spike RBD  titers ≥0.8 AU/mL |  | 236 | 269 |  | 149/236 (63.1) | 135/269 (50.2) | NS |
| Haidar, 2022 [113] | Prospective observational | - USA - Patients with autoimmune diseases | Yes | MM vs PP | Apr–Jul 2021 | Beckman Coulter SARS-CoV-2 | Anti-spike RBD IgG signal/cutoff coefficient ≥1.00 |  | 133 | 127 |  | 114/133 (85.7) | 92/127 (72.4) | NS |
| Haidar, 2022 [113] | Prospective observational | - USA - Patients with hematologic malignancies | Yes | MM vs PP | Apr–Jul 2021 | Beckman Coulter SARS-CoV-2 | Anti-spike RBD IgG signal/cutoff coefficient ≥1.00 |  | 86 | 70 |  | 46/86 (53.5) | 32/70 (45.7) | NS |
| Haidar, 2022 [113] | Prospective observational | - USA - Patients with solid tumors | Yes | MM vs PP | Apr–Jul 2021 | Beckman Coulter SARS-CoV-2 | Anti-spike RBD IgG signal/cutoff coefficient ≥1.00 |  | 76 | 60 |  | 68/76 (89.5) | 39/60 (65.0) | NS |
| Haidar, 2022 [113] | Prospective observational | - USA - Solid organ transplant recipients | Yes | MM vs PP | Apr–Jul 2021 | Beckman Coulter SARS-CoV-2 | Anti-spike RBD IgG signal/cutoff coefficient ≥1.00 |  | 217 | 228 |  | 81/217 (37.3) | 56/228 (24.6) | NS |
| Haller, 2021 [114] | Retrospective cohort | - Austria - Kidney transplant recipients | No | MM vs PP | Jan–Jun 2021 | NR | Anti-S1 RBD IgG; definition NR |  | 69 | 230 |  | 41/69 (59.4) | 102/230 (44.3) | NS |
| Hallett, 2021 [115] | Prospective cohort | - USA - Heart and lung transplant recipients | No | MM vs PP | Jan–Apr 2021 | Roche Elecsys | Anti-spike RBD  titers ≥0.8 U/mL |  | 64 | 70 |  | 41/64 (64.1) | 42/70 (60.0) | S |
| Hammer, 2022 [116] | Retrospective chart review | - Switzerland - Patients with MS | Yes | MM vs PP | NR | Abbott, Roche Elecsys | Anti-S1/S2 IgG  titers ≥100 AU/mL |  | 38 | 36 |  | 3/38 (7.9) | 2/36 (5.6) | S |
| Helfgott, 2023 [117] | Cross-sectional | - USA - Patients with AML | No | MM vs PP | NR | Roche Elecsys | Anti-spike RBD, definition NR |  | 18 | 21 |  | 14/18 (77.8) | 9/21 (42.9) | NS |
| Hirama, 2022 [118] | Open-label, nonrandomized prospective | - Japan - Lung transplant recipients | No | MM vs PP | Jun–Dec 2021 | Abbott SARS-CoV-2 IgG II Quant | Anti-S1 RBD titers ≥50 AU/mL |  | 7 | 34 |  | 3/7 (42.9) | 7/34 (20.6) | S |
| Hsu, 2022 [119] | Retrospective cohort | - USA - Patients receiving maintenance dialysis - Patients with no history of COVID-19 | No | MM vs PP | 6-month duration | ADVIA Centaur XP/XPT COV2G | Anti-S1 RBD IgG index value ≥1.0 |  | 778 | 443 |  | 645/778 (82.9) | 340/443 (76.7) | NS |
| Husain, 2021 [120] | Prospective cohort | - Country NR - Kidney transplant recipients | No | MM vs PP | NR | Roche Elecsys, DiaSorin LIAISON | Detectable anti-spike antibody |  | 12 | 16 |  | 3/12 (25.0) | 4/16 (25.0) | S |
| Jakubecz, 2022 [121] | Prospective cohort | - USA - Patients with MS and other neuroimmunologic disorders | Yes | MM vs PP | NR | Siemens Atellica IM | Anti-spike IgG  Index value ≥1.0 |  | 16 | 30 |  | 10/16 (62.5) | 16/30 (53.3) | S |
| Kappelman, 2021 [122] | Prospective cohort | - USA - Patients with IBD | No | MM vs PP | NR | LabCorp Cov2 Quant IgG | Anti-spike RBD IgG titers ≥1.0 μg/mL |  | 144 | 173 |  | 140/144 (97.2) | 159/173 (91.9) | S |
| Khan, 2022 [123] | Prospective, longitudinal cross-sectional | - USA - Patients with cancer, including solid tumors and hematologic malignancies | No | MM vs PP | NR | Roche Elecsys | Anti-spike RBD  titers ≥0.8 U/mL (≥50 AU/mL) |  | 160 | 273 |  | 101/160 (63.1) | 178/273 (65.2) | S |
| Kondo, 2022 [124] | Case control | - Country NR - Patients with inflammatory rheumatic diseases | No | MM vs PP | NR | NR | NR |  | 178 | 796 |  | 173/178 (97.2) | 663/796 (83.3) | N/A |
| Lacson, 2021 [125] | Retrospective | - USA - Patients receiving dialysis | No | MM vs PP | NR | ADVIA Centaur XP/XPT COV2G | Anti-S1 RBD titers ≥2 U/L |  | 15 | 133 |  | 14/15 (93.3) | 113/133 (85.0) | S |
| Loubet, 2022 [80] | Prospective cohort | - France - IC patients with NMOSD or MS | Yes | MMM vs PPP | Mar–Dec 2021 | EUROIMMUN ELISA | Anti-S1 IgG relative OD ≥1.1 |  | 48 | 251 |  | 40/48 (83.3) | 217/251 (86.5) | NS |
| Loubet, 2022 [80] | Prospective cohort | - France - IC patients with cancer | Yes | MMM vs PPP | Mar–Dec 2021 | EUROIMMUN ELISA | Anti-S1 IgG relative OD ≥1.1 |  | 26 | 139 |  | 26/26 (100.0) | 129/139 (92.8) | NS |
| Loubet, 2022 [80] | Prospective cohort | - France - IC patients with solid organ transplant | Yes | MMM vs PPP | Mar–Dec 2021 | EUROIMMUN ELISA | Anti-S1 IgG relative OD ≥1.1 |  | 12 | 19 |  | 5/12 (4.2) | 5/19 (26.3) | NS |
| Macrae, 2022 [126] | Prospective cohort | - Canada - Patients with cancer | No | MM vs PP | May 2021–Jul 2022 | EUROIMMUN ELISA | Anti-RBD IgG  titers ≥35.2 BAU/mL |  | 7 | 114 |  | 2/7 (28.6) | 9/114 (7.9) | S |
| Magliulo, 2022 [127] | Prospective cohort | - USA - Patients with autoimmune rheumatic disease patients | Yes | MM vs PP | Jan 2020–Feb 2021 | Siemens Atellica, ADVIA Centaur COV2G | Anti-S1 RBD index value ≥1.00 |  | 15 | 22 |  | 7/15 (46.7) | 7/22 (31.8) | S |
| Maillard, 2022 [73] | Retrospective | - France - HSCT recipients | Yes | MMM vs PPP | Jan 2021–15 Jul 2021 | Roche Elecsys, Abbott S IgG, DiaSorin TriS, Siemens, Wantai | Anti-S1 RBD titers >250 BAU/mL |  | 2 | 68 |  | 1/2 (50.0) | 28/68 (41.2) | S |
| Mairhofer, 2021 [128] | Prospective | - Austria - Patients with hematologic cancers | Yes | MM vs PP | From Mar 2021 | Abbott anti-S IgG | Anti-spike IgG  titers 5.680–7.1 BAU/mL |  | 22 | 23 |  | 14/22 (63.6) | 12/23 (52.2) | N/A |
| Manjappa, 2022 [85] | Prospective cohort | - USA - Patients with cGVHD on immune-suppression | Yes | MM vs PP | NR | Roche Elecsys | Anti-spike RBD titers ≥0.8 AU/mL |  | 7 | 24 |  | 5/7 (71.4) | 12/24 (50.0) | S |
| Mitchell, 2022 [129] | Cohort | - USA - Patients with rheumatic and musculoskeletal diseases | No | MM vs PP | Dec 2020–Jul 2021 | Roche Elecsys | Anti-spike RBD  titers ≥250 U/mL |  | 409 | 529 |  | 337/409 (82.4) | 383/529 (72.4) | NS |
| Mitchell, 2022 [129] | Cohort | - USA - Patients with solid organ transplant | No | MM vs PP | Dec 2020–Jul 2021 | Roche Elecsys | Anti-spike RBD  titers ≥250 U/mL |  | 128 | 132 |  | 99/128 (77.3) | 72/132 (54.5) | NS |
| Narasimhan, 2021 [130] | Prospective cohort | - USA - Lung transplant recipients | No | MM vs PP | Dec 2020–Mar 2021 | Abbott Alinity i | Anti-N IgG (IgGNC), anti-spike IgM (IgMSP), or anti-spike IgG (IgGSP); patients with 2 titers ≥1.4 AU/mL (IgGNC), ≥1.0 AU/mL (IgMSP), or ≥50 AU/mL (IgGSP) |  | 25 | 48 |  | 9/25 (36.0) | 9/48 (18.8) | S |
| Ollila, 2021 [131] | Retrospective cohort | - USA - Patients with hematologic malignancy | Yes | MM vs PP | Feb–Jul 2021 | Wondfo qualitative SARS-CoV-2 total antibody test, Abbott AdviseDx SARS-CoV-2 IgG II | Anti-RBD IgG/IgM; definition of seropositivity NR |  | 74 | 140 |  | 42/74 (56.8) | 50/140 (35.7) | N/A |
| Ollila, 2022 [132] | Retrospective | - USA - Patients with hematologic malignancy | Yes | MM vs PP | Feb 2021–Feb 2022 | Wondfo qualitative SARS-CoV-2 total antibody test, Abbott AdviseDx SARS-CoV-2 IgG II | Anti-RBD IgG/IgM titers ≥50 AU/mL |  | 128 | 214 |  | 77/128 (60.2) | 89/214 (41.6) | S |
| Panizo, 2022 [133] | Prospective cohort | - Spain - Patients with CKD | No | MM vs PP | Mar–Oct 2021 | Roche Elecsys | Anti-spike RBD titers ≥250 U/mL |  | 30 | 22 |  | 30/30 (100.0) | 20/22 (90.9) | NS |
| Pham, 2022 [134] | Retrospective cohort | - USA - Patients with primary immunodeficiency and functional B cell defects | No | MM vs PP | NR | ELISA | Anti-S1 RBD IgG OD ≥3 SD to mean ELISA ODs of 94 historical negative controls from healthy donors |  | 10 | 23 |  | 6/10 (60.0) | 10/23 (43.5) | NS |
| Piñana, 2022 [135] | Prospective cohort | - Spain - Patients with hematologic disorders | Yes | MM vs PP | 30 Dec 2020–30 Jun 2021 | ELISA, CLIA | SARS-CoV-2-reactive IgG titers ≥350 BAU/mL |  | 864 | 272 |  | 700/864 (81.0) | 204/272 (75.0) | S |
| Quiroga, 2022 [81] | Prospective observational cohort | - Spain - Patients with hemodialysis, kidney transplant, peritoneal dialysis, or CKD | NR | MMM vs PPP | NR | Quantitative Virclia IgG Monotest | Anti-spike IgG titers >36 U/mL |  | 462 | 162 |  | 268/462 (58.0) | 62/162 (38.3) | S |
| Rose, 2022 [136] | Prospective cohort | - USA - Patients with autoimmune rheumatic diseases treated with rituximab | Yes | MM vs PP | NR | Siemens Attelica IM COV2G, ADVIA Centaur COV2G | Anti-S1 RBD IgG index value ≥1.00 |  | 10 | 19 |  | 6/10 (60.0) | 10/19 (52.6) | S |
| Rotterdam, 2022 [137] | Prospective cohort | - USA - Patients with hematologic disorders | Yes | MM vs PP | Jul–Oct 2021 | Roche Elecsys | Anti-spike RBD titers ≥0.8 U/mL |  | 36 | 289 |  | 26/36 (72.2) | 248/289 (85.8) | NS |
| Ruddy, 2021 [138] | Prospective cohort | - USA - Patients with rheumatic and musculoskeletal diseases | Yes | MM vs PP | Jul 2020–Mar 2021 | Roche Elecsys | Anti-spike RBD titers >0.79 U/mL |  | 206 | 198 |  | 194/206 (94.2) | 184/198 (92.9) | S |
| Satyanarayan, 2022 [139] | Retrospective cohort | - USA - Patients with MS | Yes | MM vs PP | Mar–Aug 2021 | DiaSorin, Roche Elecsys,Kantaro | Detectable anti-spike RBD antibodies |  | 134 | 205 |  | 106/134 (79.1) | 140/205 (68.3) | S |
| Shah, 2022 [140] | Retrospective consecutive cohort | - Country NR - Patients with plasma cell dyscrasias | No | MM vs PP | 15 Apr–1 Jul 2021 | Roche Elecsys | Anti-spike RBD titers >100 U/mL 14 days after second dose |  | 37 | 41 |  | 28/37 (75.6) | 25/41 (61.0) | S |
| Shapiro, 2022 [141] | Cross-sectional cohort | - USA - Patients with hematologic malignancy | Yes | MM vs PP | Mar–Jul 2021 | NR | Anti-spike IgG; seropositivity definition NR |  | 36 | 70 |  | 30/36 (83.3) | 60/70 (85.7) | S |
| Sibbel, 2021 [142] | Retrospective observational | - USA - Patients receiving hemodialysis | No | MM vs PP | Jan–Mar 2021 | Diazyme Laboratories | Anti-SARS-CoV-2 IgG titers >1 AU/mL 28–56 days postvaccination |  | 321 | 315 |  | 308/321 (96.0) | 309/315 (98.1) | NS |
| Sormani, 2022 [143] | Observational multi-center prospective | - Italy - Patients with MS | Yes | MM vs PP | 4 Mar–9 Jul 2021 | Roche Elecsys | Anti-spike RBD  titers ≥0.8 U/mL |  | 23 | 131 |  | 14/23 (60.9) | 53/131 (40.5) | NS |
| Stampfer, 2021 [144] | Observational trial | - USA - Patients with multiple myeloma | No | MM vs PP | NR | ELISA | Anti-S1 RBD IgG titers >250 IU/mL |  | 48 | 48 |  | 30/48 (62.5) | 13/48 (27.1) | NS |
| Strauss, 2022 [74] | Longitudinal observational cohort | - USA - Liver transplant recipients | No | MMM vs PPP | 5 Jun–21 Dec 2021 | Roche Elecsys, EUROIMMUN ELISA | Anti-S1 RBD IgG titers ≥0.8 U/mL (Roche) and ≥1.1 AU/mL (EUROIMMUN) |  | 75 | 73 |  | 73/75 (97.3) | 65/73 (89.0) | S |
| Strauss, 2021 [145] | Prospective cohort | - USA - Post-liver transplant recipients | No | MM vs PP | 7 Jan–26 Mar 2021 | Roche Elecsys, EUROIMMUN ELISA | Anti-S1 RBD IgG titers ≥0.8 U/mL (Roche) and ≥1.1 AU/mL (EUROIMMUN) |  | 76 | 85 |  | 68/76 (89.5) | 62/85 (72.9) | S |
| Stumpf, 2021 [45] | Prospective cohort | - Germany - Patients receiving dialysis | Yes | MM vs PP | 15 Jan–24 Feb 2021 | EUROIMMUN ELISA | Anti-S1 and N IgG titers ≥1.1 AU/mL |  | 936 | 200 |  | 908/936 (97.0) | 176/200 (88.0) | S |
| Stumpf, 2021 [45] | Prospective cohort | - Germany - Kidney transplant recipients | Yes | MM vs PP | 15 Jan–24 Feb 2021 | EUROIMMUN ELISA | Anti-S1 and N IgG titers ≥1.1 AU/mL |  | 234 | 99 |  | 115/234 (49.1) | 26/99 (26.3) | S |
| Stumpf, 2022 [75] | Prospective cohort | - Germany - Kidney transplant recipients | Yes | MMM vs PPP | Jun–Dec 2021 | NR | Anti-S1 IgG titers >25.6 BAU/mL |  | 63 | 57 |  | 31/63 (49.2) | 17/57 (29.8) | NS |
| Syversen, 2022 [146] | Longitudinal observational | - Norway - Patients with weak serologic response >3 weeks after completing the standard 2-dose regimen | No | MM vs PP | 2 Feb–11 Jun 2021 | In-house assay | Anti-spike RBD IgG titers ≥70 AU/mL |  | 401 | 1,152 |  | 391/401 (97.5) | 1026/ 1152 (89.1) | NS |
| Thakkar, 2021 [147] | Retrospective cohort | - Austria - Patients with solid and hematologic cancers | Yes | MM vs PP | Mar–Jun 2021 | CLIA | Anti- S1 RBD IgG titers >50 AU/mL |  | 62 | 115 |  | 58/62 (93.5) | 109/115 (94.8) | S |
| Thompson, 2022 [76] | Retrospective cohort | - USA - Patients with hematologic malignancies | NR | MMM vs PPP | 31 Oct 2019–1 Nov 2021 | ADVIA Centaur platform | Anti-S1 RBD IgG cutoff index value >1.0 |  | 164 | 329 |  | 141/164 (86.0) | 248/329 (75.4) | S |
| Thuluvath, 2022 [148] | Prospective cohort | - USA - Patients with chronic liver disease or liver transplant recipients | No | MM vs PP | Up to 1 year | Roche semi-quantitative assay via LabCorp | Anti-trimeric S IgG titers ≥250 U/mL |  | 110 | 104 |  | 84/110 (76.4) | 67/104 (64.4) | NS |
| Tien, 2022 [149] | Prospective observational | - China - Patients with immune-mediated inflammatory diseases | Yes | MM vs PP | NR | CLIA | Anti-S1 RBD IgG cutoff index value >10 AU/mL |  | 114 | 40 |  | 98/114 (86.0) | 37/40 (92.5) | S |
| Toapanta-Yanchapaxi, 2022 [150] | Prospective observational cohort | - Mexico - Liver transplant recipients | No | MM vs PP | Feb–Sep 2021 | DiaSorin LIAISON, Abbott AdviseDx SARS-CoV-2 IgG II | Anti-spike S1/S2 IgG titers >15 AU/mL (DiaSorin) or >50 AU/mL (Abbott) |  | 4 | 65 |  | 4/4 (100.0) | 58/65 (89.2) | NS |
| Wagner, 2022 [151] | Prospective, open-label, phase IV trial | - Austria - Patients with solid tumors | No | MM vs PP | Mar–Jun 2021 | EUROIMMUN ELISA | Anti-S1 IgG  titers >35.2 BAU/mL |  | 27 | 36 |  | 27/27 (100.0) | 35/36 (97.2) | S |
| Watanabe, 2022 [77] | Prospective cohort | - Japan - Patients with hematologic malignancies receiving allogeneic HSCT | No | MMM vs PPP | Mar–Aug 2021 | QuaResearch COVID-19 human IgM IgG ELISA | Anti-S1 IgG cut-off of 0.26 |  | 7 | 15 |  | 7/7 (100.0) | 14/15 (93.3) | S |
| Werbel, 2021 [78] | Case series | - USA - Solid organ transplant recipients | No | MMM vs PPP | 20 Mar–10 May 2021 | EUROIMMUN ELISA, Roche anti-RBD pan-IgG | Anti-S1 RBD IgG titers >1.1 AU/mL (EUROIMMUN)or >0.8 U/mL (Roche) |  | 3 | 3 |  | 2/3 (66.7) | 1/3 (33.3) | S |
| Yang, 2022 [31] | Retrospective cohort | - USA - Immuno-suppressed patients | Yes | MM vs PP | 1 Jan 2021–15 Nov 2021 | EUROIMMUN ELISA | Anti-S1 RBD IgG cutoff value ≥1.1 |  | 137 | 266 |  | 89/137 (65.0) | 163/266 (61.3) | NS |
| Zacharopoulou, 2022 [152] | Prospective cohort | - Greece - Patients with IBD | No | MM vs PP | May–Aug 2021 | EUROIMMUN QuantiVac ELISA | Anti-S1 IgG  titers >11 RU/mL |  | 15 | 340 |  | 14/15 (93.3) | 336/340 (98.8) | NS |

AML, acute myeloid leukemia; AU, absorbance units; BAU, binding antibody units; BiTE, bispecific T-cell engager; CAR, chimeric antigen receptor; cGVHD, chronic graft-vs-host disease; CKD, chronic kidney disease; CLIA, chemiluminescence immunoassay; CLL, chronic lymphocytic leukemia; ELISA, enzyme-linked immunosorbent assay; HSCT, hematopoietic stem cell transplant; IBD, inflammatory bowel disease; IC, immunocompromised; Ig, immunoglobulin; IgGNC, immunoglobulin G nucleocapsid; IgGSP, immunoglobulin G spike protein; IgMSP, immunoglobulin M spike protein; mAb, monoclonal antibody; MS, multiple sclerosis; N, SARS-CoV-2 nucleocapsid; N/A, not applicable; NHL, non-Hodgkin lymphoma; NMOSD, neuromyelitis optica spectrum disorder; NR, not reported; NS, not serious; OD, optical density; RBD, receptor binding domain; RoB, risk of bias; RU, relative unit; S, serious; S1, SARS-CoV-2 spike S1 subunit; S2, SARS-CoV-2 spike S2 subunit.

MM vs PP defined as 2 doses of mRNA-1273 (100 mcg mRNA each) vs 2 doses of BNT162b2 (30 mcg mRNA each).

MMM vs PPP defined as 3 doses of mRNA-1273 (2-dose primary series of 100 mcg mRNA each followed by a booster dose of 50 mcg mRNA or 3 doses of 100 mcg mRNA each) vs 3 doses of BNT162b2 (30 mcg mRNA each).
