## Supplemental Table 4 for "Immunogenicity of mRNA-1273 and BNT162b2 in Immunocompromised Patients: Systematic Review and Meta-Analysis Using GRADE"

### Table S4. Characteristics of Studies Included in the Meta-Analysis: Total Anti-spike Binding Antibody or IgG Titers

| **Author, Year** | **Study Characteristics** | | | | | |  | **Vaccinated, n** | |  | **Total Antibody Titers** | |  | |
| --- | --- | --- | --- | --- | --- | --- | --- | --- | --- | --- | --- | --- | --- | --- |
|  | **Study Design** | **Population** | **Anti-CD20 mAb** | **Vaccination** | **Study Period** | **Total Antibody Definition** |  | **mRNA-1273** | **BNT 162b2** |  | **mRNA-1273** | **BNT 162b2** | | **RoB** |
| Abid, 2022 [66] | Retrospective cohort | - USA - Recipients of HSCT, CAR T cell, or BiTE | No | MMM vs PPP | NR | Anti-S1 RBD IgG, mean, AU/mL |  | 23 | 43 |  | 393.3 | 533.3 | | S |
| Addeo, 2021 [92] | Prospective observational cohort | - USA and Switzerland - Patients with cancer | Yes | MM vs PP | Jan–Apr 2021 | Anti-spike total binding antibody, median (IQR), U/mL |  | 93 | 30 |  | 2500.0 (442.0–2500.0) | 1232.0 (258.0–2500.0) | | NS |
| Aleman, 2021 [94] | Retrospective cohort | - USA - Patients with multiple myeloma | No | MM vs PP | NR | Anti-spike IgG, median (IQR), AU/mL |  | 37 | 99 |  | 293.96 (610.05–46.07) | 85.45 (450.45–13.09) | | S |
| Andreica, 2022 [83] | Prospective cohort | - Germany - Patients with rheumatoid arthritis, axial spondyloarthritis, or psoriatic arthritis receiving TNF inhibitor therapy | No | MM vs PP | NR | Anti-spike IgG, median (IQR), U/mL |  | 20 | 7 |  | 2477.5 (1036.0–12,477.5) | 1441.4 (225.2–2792.8) | | NS |
| Barmettler, 2022 [68] | Retrospective longitudinal cohort | - USA - Patients with predominant antibody deficiency | Yes | MMM vs PPP | Dec 2020–Jun 2021 | Anti-spike RBD total binding antibody, geometric mean (95% CI), U/mL |  | 33 | 25 |  | 305.2 (87.4, 1065.0) | 106.9 (33.9, 337.3) | | S |
| Caldera, 2022 [153] | Prospective cohort | - USA - Patients with IBD | No | MM vs PP | NR | Anti-S1 IgG, median (IQR), μg/mL |  | 71 | 81 |  | 38  (24–78) | 31  (12–56) | | NS |
| Chang, 2022 [86] | Observational cohort | - USA - Patients with NHL including CLL | Yes | MM vs PP | NR | Anti-spike IgG, median (range), AU/mL |  | 53 | 54 |  | 6586.2 (4351.72–8448.27) | 8013.79 (6337.93–9751.72) | | NS |
| Chen, 2022 [102] | Prospective observational | - Taiwan - Kidney transplant recipients | No | MM vs PP | Jul–Dec 2021 | Anti-spike total binding antibody,^a^ mean (SD), BAU/mL |  | 58 | 18 |  | 2525.7 (2229.5) | 616.9  (1128.3) | | NS |
| Dayam, 2022 [154] | Prospective cohort | - Canada - Patients with immune-mediated inflammatory diseases | No | MM vs PP | 8 Jan–4 Oct 2021 | Anti-spike trimer IgG, mean, BAU/mL |  | 8 | 105 |  | 1.596 | 1.215 | | S |
| Denault, 2022 [72] | Prospective cohort | - USA - Patients with breast cancer | No | MMM vs PPP | Apr–Aug 2021 | Anti-spike total binding antibody, geometric mean, log10(U/mL) |  | 75 | 108 |  | 3.0 | 2.6 | | NS |
| Göschl, 2022 [110] | Retrospective cohort | - Austria - Patients with inborn errors of immunity | Yes | MM vs PP | NR | Anti-S1 RBD total binding antibody, mean (SD), BAU/mL |  | 6 | 17 |  | 1173.8 (1033.82) | 780.7 (769.14) | | NS |
| Haidar, 2022 [113] | Prospective observational | - USA - Patients with autoimmune diseases | Yes | MM vs PP | Apr–Jul 2021 | Anti-S1 RBD IgG, mean (95% CI), BAU/mL |  | 133 | 127 |  | 10.34 (8.45, 12.18) | 5.19 (4.22, 6.07) | | NS |
| Haidar, 2022 [113] | Prospective observational | - USA - Patients with solid tumors | Yes | MM vs PP | Apr–Jul 2021 | Anti-S1 RBD IgG, mean (95% CI), BAU/mL |  | 76 | 60 |  | 14.21 (10.56, 17.85) | 6.87 (5.08, 8.62) | | NS |
| Haidar, 2022 [113] | Prospective observational | - USA - Patients with hematologic malignancies | Yes | MM vs PP | Apr–Jul 2021 | Anti-S1 RBD IgG, mean (95% CI), BAU/mL |  | 86 | 70 |  | 7.72 (5.88, 9.53) | 5.36 (4.04, 6.70) | | NS |
| Haidar, 2022 [113] | Prospective observational | - USA - Patients with solid organ transplant | Yes | MM vs PP | Apr–Jul 2021 | Anti-S1 RBD IgG, mean (95% CI), BAU/mL |  | 76 | 60 |  | 14.21 (10.56, 17.85) | 6.87 (5.08, 8.62) | | NS |
| Hammer, 2022 [116] | Retrospective chart review | - Switzerland - Patients with MS | Yes | MM vs PP | NR | Anti-spike IgG, geometric mean (95% CI), AU/mL |  | 38 | 36 |  | 30.2 (0.8, 59.6) | 25.6 (−1.1, 52.3) | | S |
| Helfgott, 2023 [117] | Cross-sectional | - USA - Patients with AML | No | MM vs PP | NR | Anti-spike RBD total binding antibody, median (95% CI), U/mL |  | 17 | 18 |  | 452.3  (141.73, 1398.40) | 108.5  (33.89, 1031.79) | | NS |
| Ionita, 2022 [155] | NR | - Romania - Patients receiving dialysis | No | MM vs PP | Mar–May 2021 | Anti-spike IgG, median (IQR), U/mL |  | 80 | 169 |  | 1032 (440–2687) | 297  (78–939) | | S |
| Kaiser, 2021 [156] | Retrospective chart review | - Austria - Patients with end-stage renal disease undergoing hemodialysis | Yes | MM vs PP | Feb–Mar 2021 | Anti-spike IgG, median (IQR), BAU/mL |  | 77 | 39 |  | 1507 (612–3112) | 676 (197–1363) | | N/A |
| Kappelman, 2021 [122] | Prospective cohort | - USA - Patients with IBD | No | MM vs PP | NR | Anti-spike RBD IgG, median (IQR), μg/mL |  | 144 | 173 |  | 23.95 (0–81) | 13.77 (0–50) | | S |
| Khan, 2022 [123] | Prospective, longitudinal cross-sectional | - USA - Patients with cancer, including solid tumors and hematologic malignancies | No | MM vs PP | NR | Anti-spike RBD total binding antibody, geometric mean (95% CI), U/mL |  | 160 | 273 |  | 676.07 (337.9, 1352.68) | 503.44 (318.84, 794.91) | | S |
| Kister, 2022 [157] | Observational cohort | - USA - Patients with MS | Yes | MM vs PP | Jan–Nov 2021 | Anti-spike RBD total binding antibody, geometric mean (SD), U/mL |  | 134 | 217 |  | 2.3 (1.5) | 2 (1.4) | | NS |
| Kondo, 2022 [124] | Case control | - Country NR - Patients with inflammatory rheumatic diseases | No | MM vs PP | NR | Neutralizing antibodies against SARS-CoV-2, mean (SD), IU/mL |  | 178 | 796 |  | 29.4 (33.9) | 10.8 (16.5) | | N/A |
| Long, 2022 [158] | Prospective cohort | - USA - Patients with IBD | No | MM vs PP | NR | Anti-spike IgG, median (IQR), μg/mL |  | 243 | 415 |  | 17.51 (4.4–21.0) | 39.03 (8.8–46.0) | | NS |
| Loubet, 2022 [80] | Prospective cohort | - France - IC patients with cancer | Yes | MMM vs PPP | Mar–Dec 2021 | Anti-spike IgG, geometric mean (95% CI), BAU/mL |  | 26 | 129 |  | 1899.1 (1236.1, 2917.6 | 875.2 (719.11, 1065.2) | | NS |
| Loubet, 2022 [80] | Prospective cohort | - France - IC patients with NMOSD or MS | Yes | MMM vs PPP | Mar–Dec 2021 | Anti-spike IgG, geometric mean (95% CI), BAU/mL |  | 40 | 217 |  | 1109 (710, 1732.4) | 1005.9 (854.6, 1183.9) | | NS |
| Macrae, 2022 [126] | Prospective cohort | - Canada - Patients with cancer | No | MM vs PP | May 2021–Jul 2022 | Anti-spike RBD IgG, mean (SD), BAU/mL |  | 7 | 114 |  | 1169.3 (2924.2) | 2577.7 (2906) | | S |
| Mairhofer, 2021 [128] | Prospective | - Austria - Patients with cancer | Yes | MM vs PP | From Mar 2021 | Anti-S IgG, mean (95% CI), BAU/mL |  | 43 | 44 |  | 1774.99 (1103.57, 2275.85) | 592.32 (236.04, 929.06) | | N/A |
| Manjappa, 2022 [85] | Prospective cohort | - USA - Patients with cGVHD on immune-suppression | Yes | MM vs PP | NR | Anti-spike total binding antibody, mean (range), IU/mL |  | 5 | 24 |  | 11,575 (0 – 14,621) | 2084  (70–25,000) | | S |
| Narasimhan, 2021 [130] | Prospective cohort | - USA - Lung transplant recipients | No | MM vs PP | Dec 2020–Mar 2021 | Anti-spike IgG, median (95% CI), AU/mL |  | 25 | 48 |  | 20.6 (0.8, 80.2) | 0.9  (0, 4.1) | | S |
| Nishikubo, 2022 [159] | Prospective | - Japan - Allo-HSCT recipients | Yes | MM vs PP | Jun–Sep 2021 | Anti-S1 IgG, geometric mean (95% CI), BAU/mL |  | 6 | 38 |  | 662 (10.4, 4229) | 242  (102, 572) | | NS |
| Ollila, 2022 [132] | Retrospective | - USA - Patients with hematologic malignancy | Yes | MM vs PP | Feb 2021, Feb 2022 | Anti-RBD IgG, median (IQR), AU/mL |  | 128 | 214 |  | 412.37 (13.88–2926.32) | 214.59  (1–3759.5) | | S |
| Panizo, 2022 [133] | Prospective cohort | - Spain - Patients with CKD | No | MM vs PP | Mar–Oct 2021 | Anti-spike total binding antibody, median, BAU/mL |  | 30 | 22 |  | 2500 | 381 | | NS |
| Raptis, 2022 [160] | Observational | - Switzerland - Patients with inﬂammatory rheumatic diseases | Yes | MM vs PP | 1 Mar–30 Sep 2021 | Anti-S1 IgG, median (IQR), OD ratio ≥5 |  | 228 | 267 |  | 8.2  (7.4–8.8) | 7.3  (6–8.3) | | NS |
| Shah, 2022 [140] | Retrospective consecutive cohort | - Country NR - Patients with plasma cell dyscrasias | No | MM vs PP | 15 Apr–1 Jul 2021 | Anti-spike total binding antibody 14 days after second vaccination, median (IQR), U/mL |  | 37 | 41 |  | 794.87 (0–2529) | 294.87 (0–2493) | | S |
| Shapiro, 2022 [141] | Cross-sectional cohort | - USA - Patients with hematologic malignancy | Yes | MM vs PP | Mar–Jul 2021 | Anti-spike IgG 14 days after second vaccination, median (IQR), U/mL |  | 36 | 70 |  | 5290 (50–50,000) | 1697 (50–50,000) | | S |
| Speich, 2022 [65] | Parallel, 2-arm, open-label, noninferiority RCT | - Switzerland - Solid organ transplant recipients | No | MM vs PP | 12-week follow-up | Anti-S1 RBD total binding antibody 12±1 weeks after vaccination, mean (95% CI), U/mL |  | 24 | 26 |  | 28.757 (0.2, 12.4) | 3113.6 (9.4, 36.2) | | NS^b^ |
| Stampfer, 2021 [144] | Observational trial | - USA - Patients with multiple myeloma | No | MM vs PP | NR | Anti-spike ectodomain antibody, median (IQR), U/mL |  | 48 | 48 |  | 346.2 (1.5–8215.9) | 100.6 (0.1–7715.9) | | NS |
| Stumpf, 2022 [75] | Prospective cohort | - Germany - Kidney transplant recipients | Yes | MMM vs PPP | Jun– Dec 2021 | Anti-S1 IgG, median (IQR), BAU/mL |  | 63 | 57 |  | 28.1 (3.2–209.4) | 3.8  (3.2–16.7) | | NS |
| Stumpf, 2022 [161] | Prospective observational | - Germany - Patients receiving dialysis | No | MM vs PP | Jan–Feb 2021 | Anti-S1 IgG, mean (95% CI), BAU/mL |  | 819 | 151 |  | 384 (384, 384) | 384 (194.3, 384) | | NS |
| Stumpf, 2022 [161] | Prospective observational | - Germany - Kidney transplant recipients | No | MM vs PP | Jan–Feb 2021 | Anti-S1 IgG, mean (95% CI), BAU/mL |  | 81 | 29 |  | 384 (126.9, 384) | 201.9 (90.1, 384) | | NS |
| Su, 2022 [162] | Prospective cohort | - Switzerland - Patients with metastatic solid tumors undergoing systemic treatment | No | MM vs PP | Jan–Jul 2021 | Anti-spike total binding antibody, median (IQR), BAU/mL |  | 10 | 41 |  | 910.5 (364.3–1871.8) | 219 (62.5–511.6) | | NS |
| Syversen, 2022 [146] | Longitudinal observational | - Norway - Patients with weak serologic response >3 weeks after completing the standard 2-dose regimen | No | MM vs PP | 2 Feb–11 Jun 2021 | Anti-spike RBD IgG, median (IQR), AU/mL |  | 401 | 1,152 |  | 2308 (377–8812) | 408 (170–2205) | | NS |
| Thakkar, 2021 [147] | Retrospective cohort | - Austria - Patients with solid and hematologic cancers | Yes | MM vs PP | Mar–Jun 2021 | Anti-spike IgG, mean (SD), AU/mL |  | 57 | 106 |  | 11,963 (18,742) | 5173 (16,699) | | S |
| Tien, 2022 [149] | Prospective observational | - China - Patients with immune-mediated inflammatory diseases | Yes | MM vs PP | NR | Anti-spike RBD IgG, median, AU/mL |  | 114 | 40 |  | 61.7 | 56.5 | | S |
| Toapanta-Yanchapaxi, 2022 [150] | Prospective observational cohort | - Mexico - Liver transplant recipients | No | MM vs PP | Feb–Sep 2021 | Anti-spike RBD IgG, median (IQR), AU/mL |  | 3 | 55 |  | 4071.6 (2963.7–5450.4) | 1230.9  (188.9–8658.8) | | NS |
| Vergori, 2022 [82] | Observational cohort | - Italy - Patients with HIV | No | MMM vs PPP | NR | Anti-spike RBD IgG, mean log_2_ (SD), BAU/mL |  | 80 | 44 |  | 12.4 (1.2) | 10.9 (2.6) | | NS |
| Verstappen, 2022 [163] | Prospective longitudinal cohort | - Netherlands - Patients with primary Sjögren syndrome | No | MM vs PP | As of Mar 2021 | Anti-S1 RBD IgG, median, AU/mL |  | 6 | 45 |  | 16,352 | 102,828 | | NS |
| Wagner, 2022 [151] | Prospective, open-label, phase IV trial | - Austria - Patients with multiple myeloma | No | MM vs PP | Mar–Jun 2021 | Anti-spike RBD IgG, mean (95% CI), BAU/mL |  | 22 | 48 |  | 1289 (521, 3190) | 374.8 (163.5, 828) | | S |
| Wagner, 2022 [151] | Prospective, open-label, phase IV trial | - Austria - Patients with IBD | No | MM vs PP | Mar–Jun 2021 | Anti-spike RBD IgG, mean, BAU/mL |  | 128 | 2 |  | 2703.93 | 765.36 | | S |
| Wagner, 2022 [151] | Prospective, open-label, phase IV trial | - Austria - Patients with solid tumors | No | MM vs PP | Mar–Jun 2021 | Anti-spike RBD IgG, mean (95% CI), BAU/mL |  | 27 | 36 |  | 2827 (1809, 4355) | 964.5 (582, 1595) | | S |
| Watanabe, 2022 [77] | Prospective cohort | - Japan - Patients with hematologic malignancies receiving allogeneic HSCT | No | MMM vs PPP | Mar–Aug 2021 | Anti-S1 IgG, median (range), OD |  | 7 | 14 |  | 1.28 (0.507–1.477) | 1.32 (0–1.734) | | S |
| Werbel, 2021 [78] | Case series | - USA - Solid organ transplant recipients | No | MMM vs PPP | 20 Mar–10 May 2021 | Anti–SARS-CoV-2 IgG, mean (SD), U/mL |  | 3 | 3 |  | 5.2 (4.3) | 1.4 (2) | | S |
| Zacharopoulou, 2022 [152] | Prospective cohort | - Greece - Patients with IBD | No | MM vs PP | May–Aug 2021 | Anti-S1 IgG, median (95% CI), RU/mL |  | 15 | 340 |  | 117.4 (93.3, 134.2) | 111.2 (88.1, 132.5) | | NS |

AML, acute myeloid leukemia; AU, absorbance units; BAU, binding antibody units; BiTE, bispecific T-cell engager; CAR, chimeric antigen receptor; cGVHD, chronic graft-vs-host disease; CKD, chronic kidney disease; CLL, chronic lymphocytic leukemia; HSCT, hematopoietic stem cell transplant; IBD, inflammatory bowel disease; IC, immunocompromised; Ig, immunoglobulin; IQR, interquartile range; mAb, monoclonal antibody; MS, multiple sclerosis; N, SARS-CoV-2 nucleocapsid; N/A, not applicable; NHL, non-Hodgkin lymphoma; NMOSD, neuromyelitis optica spectrum disorder; NR, not reported; NS, not serious; OD, optical density; RBD, receptor binding domain; RCT, randomized controlled trial; RoB, risk of bias; S, serious; S1, SARS-CoV-2 spike S1 subunit; S2, SARS-CoV-2 spike S2 subunit; TNF, tumor necrosis factor.

^a^Antibody titers only reported for patients who were seropositive.

^b^RCT with low RoB.

MM vs PP defined as 2 doses of mRNA-1273 (100 mcg mRNA each) versus 2 doses of BNT162b2 (30 mcg mRNA each).

MMM vs PPP defined as 3 doses of mRNA-1273 (2-dose primary series of 100 mcg mRNA each followed by a booster dose of 50 mcg mRNA or 3 doses of 100 mcg mRNA each) vs 3 doses of BNT162b2 (30 mcg mRNA each).
