## Supplemental Table 5 for "Immunogenicity of mRNA-1273 and BNT162b2 in Immunocompromised Patients: Systematic Review and Meta-Analysis Using GRADE"

### Table S5. Characteristics of Studies Included in the Meta-Analysis: Neutralizing Anti-spike Antibody Titers

| **Author, Year** | **Study Characteristics** | | | | | |  | **Vaccinated, n** | |  | **Neutralizing Antibody Titers** | |  |
| --- | --- | --- | --- | --- | --- | --- | --- | --- | --- | --- | --- | --- | --- |
|  | **Study Design** | **Population** | **Anti-CD20 mAb** | **Vaccination** | **Study Period** | **Neutralizing Antibody Definition** |  | **mRNA-1273** | **BNT 162b2** |  | **mRNA-1273** | **BNT 162b2** | **RoB** |
| Andreica, 2022 [83] | Prospective cohort | - Germany - Patients with rheumatoid arthritis, axial spondyloarthritis, or psoriatic arthritis receiving TNF inhibitor therapy | No | MM vs PP | NR | ND_50_, median (IQR), U/mL |  | 18 | 7 |  | 300.0 (37.5–1,087.5) | 206.3 (75.0–675.0) | NS |
| Chang, 2022 [86] | Observational cohort | - USA - Patients with NHL including CLL | Yes | MM vs PP | NR | ND_50_, median (IQR) |  | 53 | 54 |  | 820.08 (634.00–930.76) | 812.08 (574.72–930.76) | NS |
| Garcia-Cirera, 2022 [84] | Cross-sectional | - Spain - Patients with SLE | Yes | MM vs PP | NR | Anti–SARS-CoV-2–specific neutralizing antibody titer, mean (SD) |  | 27 | 12 |  | 1614 (2707.1) | 2141.8 (4280.9) | S |
| Loubet, 2022 [80] | Prospective cohort | - France - IC patients with NMOSD or MS | Yes | MMM vs PPP | Mar–Dec 2021 | Anti–SARS-CoV-2 neutralizing antibody titer, geometric mean (95% CI), BAU/mL |  | 38 | 204 |  | 370.3 (227.8, 602.0) | 203.7 (171.1, 241.6) | NS |
| Manjappa, 2022 [85] | Prospective cohort | - USA - Patients with cGVHD on immune-suppression | Yes | MM vs PP | NR | ND_50_, mean (range), IU/mL |  | 7 | 24 |  | 0.30 (0–1.45) | 1.62 (0.05–4.56) | S |
| Vergori, 2022 [82] | Observational cohort | - Italy - Patients with HIV | NR | MMM vs PPP | NR | Change of neutralizing antibody titer, mean log_2_ (SD) |  | 80 | 44 |  | 9 (1.9) | 8.3 (2.3) | NS |
| Zeng, 2022 [87] | Observational | - USA - Patients with cancer | No | MM vs PP | NR | NT_50_, mean (IQR) |  | 12 | 11 |  | 3037 (2033.18–4045.0) | 1418 (572.61–2282.92) | N/A |

AU, absorbance units; BAU, binding antibody units; cGVHD, chronic graft-vs-host disease; CLL, chronic lymphocytic leukemia; IC, immunocompromised; IQR, interquartile range; mAb, monoclonal antibody; MS, multiple sclerosis; N/A, not applicable; ND_50_, half-maximal neutralizing antibody concentration; NHL, non-Hodgkin lymphoma; NMOSD, neuromyelitis optica spectrum disorder; NR, not reported; NS, not serious; NT_50_, 50% neutralization titer; RoB, risk of bias; S, serious; SLE, systemic lupus erythematosus; TNF, tumor necrosis factor.

MM vs PP defined as 2 doses of mRNA-1273 (100 mcg mRNA each) vs 2 doses of BNT162b2 (30 mcg mRNA each).

MMM vs PPP defined as 3 doses of mRNA-1273 (2-dose primary series of 100 mcg mRNA each followed by 50 mcg mRNA booster dose or 3 doses of 100 mcg RNA each) vs 3 doses of BNT162b2 (30 mcg mRNA each).
