## Supplemental Table 6 for "Immunogenicity of mRNA-1273 and BNT162b2 in Immunocompromised Patients: Systematic Review and Meta-Analysis Using GRADE"

### Table S6. Characteristics of Studies Included in the Meta-Analysis: Cellular Immune Response

| **Author, Year** | **Study Characteristics** | | | | | | |  | **Vaccinated, n** | |  | **Cellular Immunity** | |  |
| --- | --- | --- | --- | --- | --- | --- | --- | --- | --- | --- | --- | --- | --- | --- |
|  | **Study Design** | **Population** | **Anti-CD20 mAb** | **Vaccination** | **Study Period** | **Assay/ Platform** | **Cellular Immunity Definition** |  | **mRNA-1273** | **BNT 162b2** |  | **mRNA-1273** | **BNT 162b2** | **RoB** |
| Alfonso-Dunn, 2022 [164] | Prospective cohort | - USA - Patients with MS | Yes | MM vs PP | Jan–Nov 2021 | Immunospot Fluoro Spot | IFN-γ+/IL-2+ T cells, mean (95% CI), SFU/10^6^ PBMCs |  | 18 | 22 |  | 110.6 (72.4, 155.9) | 143.8 (94.1, 211.1) | NS |
| Caldera, 2022 [153] | Prospective cohort | - USA - Patients with IBD | No | MM vs PP | NR | Mabtech human IFN-γ Fluorospot^Plus^ | Postvaccination antigen-specific T-cell response, median (IQR), μg/mL |  | 69 | 82 |  | 352 (120–1008) | 380 (146–1377) | NS |
| Chen, 2022 [102] | Prospective observational | - Taiwan - Kidney transplant recipients | No | MM vs PP | Jul–Dec 2021 | EUROIMMUN Quan-T-cell SARS-CoV-2 IGRA | IFN-γ,^a^ mean (SD), mIU/mL |  | 37 | 14 |  | 1460.30 (931.19) | 746.54 (819.49) | NS |
| Chung, 2022 [71] | Prospective cohort | - USA - Patients with multiple myeloma or plasma cell dyscrasias | No | MMM vs PPP | Jan–Dec 2021 | Adaptive Biotech-nologies immunoSEQ T-MAP COVID | Spike-specific T-cell breadth (proportion of unique spike-specific TCR divided by the number of unique TCRs, median (IQR) |  | 22 | 28 |  | 4.63×10^−5^ (2.45×10^−6^ – 7.87×10^−5^) | 2.46×10^−5^ (8.5×10^−6^–4.1×10^−5^) | S |
| Dayam, 2022 [154] | Prospective cohort | - Canada - Patients with immune-mediated inflammatory diseases | No | MM vs PP | 8 Jan–4 Oct 2021 | BioLegend LEGENDplex CD8/NK multiplex cytokine bead assay | Spike-specific T-cell cytokine response, median (log change from background, pg/mL) |  | 8 | 97 |  | 77.11 | 58.09 | S |
| Greenberger, 2022 [112] | Prospective cohort registry | - USA - Patients with hematologic malignancies | Yes | MM vs PP | May–Dec 2021 | Adaptive Biotechnologies ImmunoSEQ T DETECTCOVID | Postvaccination spike-specific T-cell response, median (IQR), AU/mL |  | 236 | 269 |  | 10.04 (7.92–13.03) | 11.27 (8.80–15.49) | NS |
| Kister, 2022 [157] | Observational cohort | - USA - Patients with MS | Yes | MM vs PP | Jan–Nov 2021 | Rules-Based Medicine TruCulture; Thermo Fisher IFN-γ quantitative ELISA | IFN-γ, mean (SD) |  | 134 | 217 |  | 1.9 (1) | 1.5 (1) | NS |
| Kondo, 2022 [124] | Case control | - Country NR - Patients with inflammatory rheumatic diseases | No | MM vs PP | NR | IGRA | IFN-γ for antigen 2, mean (SD), IU/mL |  | 178 | 796 |  | 1.4 (1.9) | 1.0 (2.1) | N/A |
| Mairhofer, 2021 [128] | Prospective | - Austria - Patients with hematologic cancers | Yes | MM vs PP | From Mar 2021 | Miltenyi Biotec SARS-CoV-2 Prot_S human T-cell analysis kit | Spike-activated CD4^+^ T cell, mean (95% CI), % |  | 41 | 42 |  | 0.33 (0.22, 0.46) | 0.18 (0.11, 0.26) | N/A |
| Panizo, 2022 [133] | Prospective cohort | - Spain - Patients with CKD | No | MM vs PP | Mar–Oct 2021 | BD FastImmune flow cytometry | Anti-spike IFN-γ^+^ CD4^+^ T cell, median (range), % |  | 30 | 22 |  | 0.04 (0.2–2.64) | 0.02 (0–4.08) | NS |
| Stumpf, 2022 [75] | Prospective cohort | - Germany - Kidney transplant recipients | Yes | MMM vs PPP | Jun–Dec 2021 | IGRA | IFN-γ, median (IQR), mIU/mL |  | 16 | 16 |  | 28.6 (6.1–200.4) | 8.3 (1.3–60.3) | NS |
| Su, 2022 [162] | Prospective cohort | - Switzerland - Patients with metastatic solid tumors undergoing systemic treatment | Yes | MM vs PP | Jan–Jul 2021 | ELISpot | SARS-CoV-2–specific IFN-γ^+^ T cells, median (IQR), SFU per 10^6^ PBMCs |  | 10 | 41 |  | 345.6 (280.1–591.8) | 239.4 (63.2–402.1) | NS |
| Vergori, 2022 [82] | Observational cohort | - Italy - Patients with HIV | NR | MMM vs PPP | NR | ELISA | Change in IFN-γ from second to third dose, mean log_2_ (SD), pg/mL |  | 77 | 41 |  | 7.9 (2.2) | 6.5 (3.3) | NS |
| Verstappen, 2022 [163] | Prospective longitudinal cohort | - Netherlands - Patients with primary Sjögren syndrome | No | MM vs PP | As of Mar 2021 | ELISpot | Change in spike-specific IFN-γ–producing SFCs per 10^6^ PBMCs |  | 2 | 18 |  | 833 | 338 | NS |

BAU, binding antibody units; CKD, chronic kidney disease; ELISA, enzyme-linked immunosorbent assay; ELISpot, enzyme-linked immunosorbent spot; IBD, inflammatory bowel disease; IFN, interferon; IGRA, interferon gamma release assay; IL, interleukin; IQR, interquartile range; IU, international unit; mAb, monoclonal antibody; MS, multiple sclerosis; N/A, not applicable; NR, not reported; NS, not serious; PBMC, peripheral bone mononuclear cell; RoB, risk of bias; S, serious; SFC, spot-forming cell; SFU, spot-forming unit; TCR, T-cell receptor.

^a^IFN-γ concentrations reported only for patients with a positive cellular immune response.

MM vs PP defined as 2 doses of mRNA-1273 (100 mcg mRNA each) vs 2 doses of BNT162b2 (30 mcg mRNA each).

MMM vs PPP defined as 3 doses of mRNA-1273 (2-dose primary series of 100 mcg mRNA each followed by 50 mcg mRNA booster dose or 3 doses of 100 mcg mRNA each) vs 3 doses of BNT162b2 (30 mcg mRNA each).
