## Supplemental Table 7 for "Immunogenicity of mRNA-1273 and BNT162b2 in Immunocompromised Patients: Systematic Review and Meta-Analysis Using GRADE"

### Table S7. RoB Assessment per the Newcastle-Ottawa Scale for Cohort and Cross-Sectional Studies

| **I. Cohort Studies** | | | | | |  | | | | |
| --- | --- | --- | --- | --- | --- | --- | --- | --- | --- | --- |
| **Author, Year** | **Total Score** | **Stars Given, n** | | | | | | | | |
|  |  | **Representativeness of Exposed Cohort^a^** | **Selection of Nonexposed Cohort^b^** | **Ascertainment of Exposure^c^** | **Outcome Not Present at Baseline^d^** | | **Comparability of Cohorts^e^** | **Assessment of Outcome^f^** | **Sufficient Follow-Up Duration^g^** | **Adequate Follow-Up^h^** |
| Abid, 2022 [66] | 6 stars | 1 | 0 | 1 | 1 | | 0 | 1 | 1 | 1 |
| Addeo, 2021 [92] | 7 stars | 1 | 1 | 1 | 1 | | 0 | 1 | 1 | 1 |
| Agha, 2021 [93] | 6 stars | 1 | 0 | 1 | 1 | | 0 | 1 | 1 | 1 |
| Aleman, 2021 [94] | 5 stars | 0 | 0 | 1 | 1 | | 0 | 1 | 1 | 1 |
| Alfonso-Dunn, 2022 [164] | 7 stars | 1 | 1 | 1 | 1 | | 0 | 1 | 1 | 1 |
| Anand, 2021 [95] | 7 stars | 1 | 1 | 1 | 1 | | 0 | 1 | 1 | 1 |
| Andreica, 2022 [83] | 8 stars | 1 | 1 | 1 | 1 | | 1 | 1 | 1 | 1 |
| Arranz, 2022 [96] | 6 stars | 1 | 0 | 0 | 1 | | 1 | 1 | 1 | 1 |
| Arroyo-Sánchez, 2022 [97] | 7 stars | 1 | 1 | 1 | 1 | | 0 | 1 | 1 | 1 |
| Arumahandi de Silva, 2022 [98] | 7 stars | 1 | 0 | 1 | 1 | | 1 | 1 | 1 | 1 |
| Bae, 2022 [99] | 8 stars | 1 | 1 | 1 | 1 | | 1 | 1 | 1 | 1 |
| Bagacean, 2022 [67] | 4 stars | 1 | 0 | 1 | 1 | | 0 | 0 | 1 | 0 |
| Barmettler, 2022 [68] | 6 stars | 1 | 0 | 1 | 0 | | 1 | 1 | 1 | 1 |
| Broseta, 2021 [101] | 8 stars | 1 | 1 | 1 | 1 | | 1 | 1 | 1 | 1 |
| Caldera, 2022 [153] | 8 stars | 1 | 1 | 1 | 1 | | 1 | 1 | 1 | 1 |
| Chaekal, 2022 [69] | 7 stars | 1 | 1 | 1 | 1 | | 1 | 1 | 0 | 1 |
| Chang, 2022 [86] | 8 stars | 1 | 1 | 1 | 1 | | 1 | 1 | 1 | 1 |
| Chen, 2022 [102] | 7 stars | 1 | 1 | 1 | 1 | | 0 | 1 | 1 | 1 |
| Chiang, 2022 [70] | 8 stars | 1 | 1 | 1 | 1 | | 1 | 1 | 1 | 1 |
| Chung, 2022 [71] | 6 stars | 1 | 1 | 1 | 0 | | 0 | 1 | 1 | 1 |
| Dayam, 2022 [154] | 5 stars | 1 | 1 | 1 | 0 | | 0 | 1 | 1 | 0 |
| Debska-Slizien, 2022 [104] | 8 stars | 1 | 1 | 1 | 1 | | 1 | 1 | 1 | 1 |
| Denault, 2022 [72] | 7 stars | 0 | 1 | 1 | 1 | | 1 | 1 | 1 | 1 |
| Doukas, 2022 [105] | 4 stars | 1 | 0 | 0 | 0 | | 1 | 1 | 1 | 0 |
| Ferri, 2021 [106] | 6 stars | 1 | 1 | 1 | 0 | | 0 | 1 | 1 | 1 |
| Floyd, 2022 [107] | 7 stars | 1 | 1 | 1 | 1 | | 0 | 1 | 1 | 1 |
| Fujieda, 2022 [108] | 5 stars | 1 | 1 | 1 | 1 | | 0 | 1 | 0 | 0 |
| Gallais, 2022 [79] | 7 stars | 0 | 1 | 1 | 1 | | 1 | 1 | 1 | 1 |
| Giannella, 2022 [109] | 7 stars | 0 | 1 | 1 | 1 | | 1 | 1 | 1 | 1 |
| Göschl, 2022 [110] | 8 stars | 1 | 1 | 1 | 1 | | 1 | 1 | 1 | 1 |
| Greenberger, 2021 [111] | 6 stars | 1 | 0 | 1 | 1 | | 0 | 1 | 1 | 1 |
| Greenberger, 2022 [112] | 8 stars | 1 | 1 | 1 | 1 | | 1 | 1 | 1 | 1 |
| Haidar, 2022 [113] | 8 stars | 1 | 1 | 1 | 1 | | 1 | 1 | 1 | 1 |
| Haller, 2021 [114] | 7 stars | 1 | 0 | 1 | 1 | | 1 | 1 | 1 | 1 |
| Hallett, 2021 [115] | 6 stars | 1 | 0 | 1 | 1 | | 0 | 1 | 1 | 1 |
| Hammer, 2022 [116] | 6 stars | 0 | 1 | 1 | 1 | | 0 | 1 | 1 | 1 |
| Hirama, 2022 [118] | 6 stars | 0 | 1 | 1 | 1 | | 0 | 1 | 1 | 1 |
| Hsu, 2022 [119] | 8 stars | 1 | 1 | 1 | 1 | | 1 | 1 | 1 | 1 |
| Husain, 2021 [120] | 5 stars | 1 | 0 | 1 | 1 | | 0 | 1 | 0 | 1 |
| Ionita, 2022 [155] | 6 stars | 1 | 1 | 1 | 1 | | 1 | 1 | 0 | 0 |
| Jakubecz, 2022 [121] | 5 stars | 1 | 1 | 1 | 0 | | 0 | 1 | 0 | 1 |
| Kappelman, 2021 [122] | 6 stars | 1 | 0 | 1 | 1 | | 0 | 1 | 1 | 1 |
| Khan, 2022 [123] | 6 stars | 1 | 0 | 1 | 1 | | 0 | 1 | 1 | 1 |
| Kister, 2022 [157] | 9 stars | 1 | 1 | 1 | 1 | | 2 | 1 | 1 | 1 |
| Lacson, 2021 [125] | 5 stars | 1 | 0 | 1 | 1 | | 0 | 1 | 0 | 1 |
| Long, 2022 [158] | 7 stars | 1 | 1 | 0 | 1 | | 1 | 1 | 1 | 1 |
| Loubet, 2022 [80] | 7 stars | 1 | 1 | 1 | 1 | | 1 | 1 | 0 | 1 |
| Macrae, 2022 [126] | 6 stars | 1 | 0 | 0 | 1 | | 1 | 1 | 1 | 1 |
| Magliulo, 2022 [127] | 6 stars | 1 | 0 | 1 | 1 | | 0 | 1 | 1 | 1 |
| Maillard, 2022 [73] | 6 stars | 1 | 0 | 1 | 1 | | 0 | 1 | 1 | 1 |
| Manjappa, 2022 [85] | 6 stars | 1 | 1 | 1 | 1 | | 0 | 1 | 1 | 0 |
| Mitchell, 2022 [129] | 7 stars | 0 | 1 | 1 | 1 | | 1 | 1 | 1 | 1 |
| Narasimhan, 2021 [130] | 6 stars | 1 | 1 | 1 | 0 | | 0 | 1 | 1 | 1 |
| Nishikubo, 2022 [159] | 7 stars | 1 | 1 | 1 | 1 | | 1 | 1 | 1 | 0 |
| Ollila, 2022 [132] | 6 stars | 1 | 0 | 1 | 1 | | 0 | 1 | 1 | 1 |
| Panizo, 2022 [133] | 8 stars | 1 | 1 | 1 | 1 | | 1 | 1 | 1 | 1 |
| Pham, 2022 [134] | 7 stars | 1 | 1 | 1 | 1 | | 0 | 1 | 1 | 1 |
| Piñana, 2022 [135] | 5 stars | 1 | 1 | 1 | 0 | | 0 | 1 | 1 | 0 |
| Quiroga, 2022 [81] | 5 stars | 1 | 1 | 1 | 0 | | 0 | 1 | 1 | 0 |
| Raptis, 2022 [160] | 8 stars | 1 | 1 | 1 | 1 | | 1 | 1 | 1 | 1 |
| Rose, 2022 [136] | 6 stars | 1 | 0 | 1 | 1 | | 0 | 1 | 1 | 1 |
| Rotterdam, 2022 [137] | 7 stars | 0 | 1 | 1 | 1 | | 1 | 1 | 1 | 1 |
| Ruddy, 2021 [138] | 5 stars | 1 | 0 | 1 | 0 | | 0 | 1 | 1 | 1 |
| Satyanarayan, 2022 [139] | 6 stars | 1 | 0 | 1 | 1 | | 1 | 1 | 0 | 1 |
| Shah, 2022 [140] | 6 stars | 1 | 0 | 1 | 1 | | 0 | 1 | 1 | 1 |
| Shapiro, 2022 [141] | 6 stars | 1 | 0 | 1 | 1 | | 0 | 1 | 1 | 1 |
| Sibbel, 2021 [142] | 8 stars | 1 | 1 | 1 | 1 | | 1 | 1 | 1 | 1 |
| Sormani, 2022 [143] | 7 stars | 1 | 1 | 1 | 1 | | 1 | 1 | 1 | 0 |
| Stampfer, 2021 [144] | 7 stars | 1 | 1 | 1 | 1 | | 1 | 1 | 0 | 1 |
| Strauss, 2022 [74] | 3 stars | 0 | 0 | 0 | 1 | | 1 | 0 | 1 | 0 |
| Strauss, 2021 [145] | 5 stars | 1 | 1 | 1 | 1 | | 1 | 0 | 0 | 0 |
| Stumpf, 2021 [45] | 6 stars | 1 | 1 | 1 | 0 | | 0 | 1 | 1 | 1 |
| Stumpf, 2022 [75] | 8 stars | 1 | 1 | 1 | 1 | | 1 | 1 | 1 | 1 |
| Stumpf, 2022 [161] | 8 stars | 1 | 1 | 1 | 1 | | 1 | 1 | 1 | 1 |
| Su, 2022 [162] | 7 stars | 0 | 1 | 1 | 1 | | 1 | 1 | 1 | 1 |
| Syversen, 2022 [146] | 7 stars | 1 | 0 | 1 | 1 | | 1 | 1 | 1 | 1 |
| Thakkar, 2021 [147] | 5 stars | 1 | 1 | 1 | 1 | | 0 | 1 | 0 | 0 |
| Thompson, 2022 [76] | 5 stars | 1 | 1 | 1 | 1 | | 0 | 1 | 0 | 0 |
| Thuluvath, 2022 [148] | 8 stars | 1 | 1 | 1 | 1 | | 1 | 1 | 1 | 1 |
| Tien, 2022 [149] | 6 stars | 1 | 1 | 1 | 1 | | 0 | 1 | 1 | 0 |
| Toapanta-Yanchapaxi, 2022 [150] | 8 stars | 1 | 1 | 1 | 1 | | 1 | 1 | 1 | 1 |
| Vergori, 2022 [82] | 8 stars | 1 | 1 | 1 | 1 | | 1 | 1 | 1 | 1 |
| Verstappen, 2022 [163] | 8 stars | 1 | 1 | 1 | 1 | | 1 | 1 | 1 | 1 |
| Wagner, 2022 [151] | 6 stars | 0 | 1 | 1 | 1 | | 0 | 1 | 1 | 1 |
| Watanabe, 2022 [77] | 6 stars | 1 | 1 | 1 | 0 | | 0 | 1 | 1 | 1 |
| Werbel, 2021 [78] | 4 stars | 1 | 1 | 0 | 0 | | 0 | 1 | 0 | 1 |
| Yang, 2022 [31] | 7 stars | 1 | 1 | 1 | 1 | | 0 | 1 | 1 | 1 |
| Zacharopoulou, 2022 [152] | 7 stars | 1 | 1 | 1 | 0 | | 1 | 1 | 1 | 1 |
| **II. Cross-Sectional Studies** | |  |  |  |  | |  |  |  |  |
| **Author, Year** | **Total Score** | **Stars Given, n** | | | | | | | |  |
|  |  | **Representativeness of Exposed Cohort^a^** | **Sample Size^i^** | **Non-Respondents^j^** | **Ascertainment of Exposure^k^** | | **Comparability^l^** | **Assessment of Outcome^m^** | **Statistical Test^n^** |  |
| Garcia-Cirera, 2022 [84] | 6 stars | 1 | 0 | 1 | 1 | | 1 | 1 | 1 |  |
| Helfgott, 2023 [117] | 7 stars | 1 | 1 | 1 | 1 | | 1 | 1 | 1 |  |
| Shapiro, 2022 [141] | 5 stars | 1 | 0 | 0 | 1 | | 1 | 1 | 1 |  |

IC, immunocompromised; RoB, risk of bias.

^a^1 star was given if the study population was truly or somewhat representative of a community or population. No star was given if the study population was sampled from a special population (eg, hospitalized patients).

^b^1 star was given if the nonexposed cohort (ie, non-IC cohort) was drawn from the same population as the exposed cohort (ie, IC cohort). If only 1 cohort of patients was included, no star was given.

^c^1 star was given if secured medical records or a structured interview was used to ascertain the IC condition. No star was given if the IC condition was self-reported or not described.

^d^1 star was given if the outcomes were assessed at the beginning of the study. No star was given if outcomes were not assessed at the beginning of the study.

^e^2 stars were given if the study was adjusted for the most important factors deliberately. 1 star was given if the study was adjusted for other important factors. If no adjustment was performed or there was no description of comparability, no star was given.

^f^1 star was given if the outcome was assessed from medical records or record linkage. No star was given if the outcome was self-reported.

^g^1 star was given if the duration of follow-up was >1 month. Otherwise, no star was given.

^h^1 star was given if there was complete follow-up or the lost to follow-up rate was ≤20%. No star was given if the follow-up rate was <80% or if the follow-up rate was not reported.

^i^1 star was given if the sample size was justified in the study and satisfactory. Otherwise, no star was given.

^j^1 star was given if comparability between characteristics of respondents (eg, patients who seroconverted after vaccination) and non-respondents (eg, patients who did not experience seroconversion after vaccination) was established, and if response rate was satisfactory. If the response rate or if the comparability between respondents and nonrespondents was not described or unsatisfactory, no stars were given.

^k^If a validated screening or surveillance tool was used, 2 stars were given. 1 star was given if a nonvalidated screening or surveillance tool was used but the tool is available or described. No stars were given if the measurement tool was not described or reported.

^l^If potential confounders were investigated, 1 star was given. Otherwise, no stars were given.

^m^If outcomes were assessed through independent, blind assessment or record linkage, 2 stars were given. 1 star was given if the outcome was self-reported. No stars were given if outcome assessments were not described or reported.

^n^1 star was given if the statistical test used to analyze the data was clearly described and appropriate. Otherwise, no stars were given.
