## Supplemental Table 8 for "Immunogenicity of mRNA-1273 and BNT162b2 in Immunocompromised Patients: Systematic Review and Meta-Analysis Using GRADE"

### Table S8. RoB Assessment for RCTs

| *●* Study**:** Speich B *et al.* *Clin. Infect. Dis.* 75(1), e585–e593 (2022) [65] | |
| --- | --- |
| Domain 1: RoB Arising From the Randomization Process |  |
| 1.1 Was the allocation sequence random? | Yes |
| 1.2 Was the allocation sequence concealed until participants were enrolled and assigned to interventions? | No information |
| 1.3 Did baseline differences between intervention groups suggest a problem with the randomization process? | No |
| 1.0 Algorithm result / assessor’s judgement | Low / low |
| Domain 2: RoB Due to Deviations From the Intended Interventions (Effect of Assignment to Intervention) |  |
| 2.1 Were participants aware of their assigned intervention status during the trial? | Yes |
| 2.2 Were carers and people delivering the interventions aware of the participants’ assigned intervention during the trial? | Yes |
| 2.3 If yes, probably yes, or no information to 2.1 or 2.2, were there deviations from the intended intervention that arose because of the trial context? | No |
| 2.4 If yes or probably yes to 2.3, were these deviations likely to have affected the outcome? | N/A |
| 2.5 If yes or probably yes to 2.4, were these deviations from intended intervention balanced between groups? | N/A |
| 2.6 Was an appropriate analysis used to estimate the effect of assignment to intervention? | Yes |
| 2.7 If no, probably no, or no information to 2.6, was there potential for a substantial impact on the result of the failure to analyze participants in the group to which they were randomized? | N/A |
| 2.0 Algorithm result / assessor’s judgement | Low / low |
| Domain 2: RoB Due to Deviations From the Intended Interventions (Effect of Adhering to Intervention) |  |
| 2.1 Were participants aware of their assigned intervention status during the trial? | Yes |
| 2.2 Were carers and people delivering the interventions aware of the participants’ assigned intervention during the trial? | Yes |
| 2.3 If yes, probably yes, or no information to 2.1 or 2.2, were important nonprotocol interventions balanced across intervention groups? | Yes |
| 2.4 If applicable, were there failures in implementing the intervention that could have affected the outcome? | No |
| 25. If applicable, was there nonadherence to the assigned intervention regimen that could have affected participants’ outcomes? | Probably no |
| 2.6 If no, probably no, or no information to 2.3 or yes, probably yes, or no information to 2.4 or 2.5, was an appropriate analysis used to estimate the effect of adhering to the intervention? | N/A |
| 2.0 Algorithm result / assessor’s judgement | Low / low |
| Domain 3: Missing Outcome Data |  |
| 3.1 Were data for this outcome available for all, or nearly all, participants randomized? | Yes |
| 3.2 If no, probably no, or no information to 3.1, is there evidence that the result was not biased by missing outcome data? | N/A |
| 3.3 If no or probably no to 3.2, could missingness in the outcome depend on its true value? | N/A |
| 3.4 If yes, probably yes, or no information to 3.3, is it likely that missingness in the outcome depended on its true value? | N/A |
| 3.0 Algorithm result / assessor’s judgement | Low / low |
| Domain 4: Measurement of the Outcome |  |
| 4.1 Was the method of measuring the outcome appropriate? | No |
| 4.2 Could measurement or ascertainment of the outcome have differed between intervention groups? | No |
| 4.3 If no, probably no, or no information to 4.1 and 4.2, were outcome assessors aware of the intervention received by the participants? | Yes |
| 4.4 If yes, probably yes, or no information to 4.3, could assessment of the outcome have been influenced by knowledge of the intervention received? | No |
| 4.5 If yes, probably yes, or no information to 4.4, is it likely that assessment of the outcome was influenced by knowledge of the intervention received? | N/A |
| 4.0 Algorithm result / assessor’s judgement | Low / low |
| Domain 5: Selection of the Reported Result |  |
| 5.1 Were the data that produced this result analyzed in accordance with a prespecified analysis plan that was finalized before unblinded outcome data were available for analysis? | Yes |
| 5.2 Is the numerical result being assessed likely to have been selected, on the basis of the results, from multiple eligible outcome measurements (eg, scales, definitions, time points) within the outcome domain? | No |
| 5.3 Is the numerical result being assessed likely to have been selected, on the basis of the results, from multiple eligible analyses of the data? | Probably no |
| 5.0 Algorithm result / assessor’s judgement | Low / low |
| Domain 6: Overall Bias |  |
| 6.0 Algorithm result / assessor’s judgement | Low / low |

N/A, not applicable; RCT, randomized controlled trial; RoB, risk of bias.
