## Supplemental Table 9 for "Immunogenicity of mRNA-1273 and BNT162b2 in Immunocompromised Patients: Systematic Review and Meta-Analysis Using GRADE"

### Table S9. GRADE Summary of Findings for Seroconversion Overall, by Disease Subgroup, and by Anti-CD20 Treatment

|  | **Certainty Assessment** | | | | | | | **Vaccinated With mRNA-1273,  n/N (%)** | **Vaccinated With BNT162b2, n/N (%)** | **RR  (95% CI)** | **Absolute Effect (95% CI)** | **Certainty** | **Importance** |
| --- | --- | --- | --- | --- | --- | --- | --- | --- | --- | --- | --- | --- | --- |
|  | **Studies, n** | **Study Design** | **RoB** | **Inconsistency** | **Indirect-ness** | **Imprecision** | **Other Considerations** |  |  |  |  |  |  |
| **Overall** |  | | | | | | | | | | | | |
| **Anti-CD20 mAb treatment** | 36 | nR | NS | Serious^a^ | Serious^b^ | Very serious^c^ | Strong association | 3468/4376  (79.3) | 3411/5029  (67.8) | 1.08 (1.04, 1.13) | 5933.32 more per 100,000 (3302.54, 8564.1) | Type 4^d^ | Critical |
| **No anti-CD20 mAb treatment** | 39 | nR | NS | NS^e^ | Serious^b^ | Very serious^f^ | Strong association | 3414/4334 (78.8) | 5261/7522 (69.9) | 1.10 (1.08, 1.13) | 9677.66 more per 100,000 (7515.61, 11,839.72) | Type3^g^ | Critical |
| **Autoimmune disease** |  | | | | | | | | | | | | |
| **Anti-CD20 mAb treatment** | 13 | nR | NS | NS^h^ | Serious^b^ | Very serious^i^ | None | 645/806 (80.0) | 1207/1609 (75.0) | 1.05 (0.99, 1.11) | 3922.05 more per 100,000 (−38.2, 7882.29) | Type3^g^ | Critical |
| **No anti-CD20 mAb treatment** | 7 | nR | NS | NS^j^ | Serious^b^ | NS | none | 1079/1173 (92.0) | 2736/3181 (86.0) | 1.09 (1.05, 1.14) | 8024.22 more per 100,000 (4407.81, 11,640.64) | Type3^k^ | Critical |
| **Hematologic malignancy** |  | | | | | | | | | | | | |
| **Anti-CD20 mAb treatment** | 16 | nR | NS | Serious^m^ | Serious^b^ | NS | Strong association | 1771/2413  (73.4) | 1798/2882  (62.4) | 1.15 (1.06, 1.24) | 8469.2 more per 100,000 (3918.87, 13,019.53) | Type 3^n^ | Critical |
| **No anti-CD20 mAb treatment** | 8 | nR | NS | Serious^o^ | Serious^b^ | NS | Strong association | 169/226 (74.8) | 225/362 (62.2) | 1.25 (1.07, 1.46) | 16,398.71 more per 100,000 (6904.68, 25,892.74) | Type 3^n^ | Critical |

GRADE, Grading of Recommendations, Assessment, Development, and Evaluation; mAb, monoclonal antibody; nR, nonrandomized; NS, not serious; RoB, risk of bias; RR, risk ratio.

^a^*I*²=48.4%, Χ²=67.76, p(Q)=0; moderate heterogeneity.

^b^Outcome definitions rather heterogeneous (different antibody types and assays used).

^c^Wider 95% CI due to only 1 event in the mRNA-1273 arm in Gallais 2022 and Maillard 2022 and the overall small sample size of Werbel 2021.

^d^Lower grading due to imprecision and indirectness because of varying outcome definitions (different antibody types and assays used).

^e^*I*²=42.7%, Χ²=66.31, p(Q)=0; moderate heterogeneity.

^f^Wider 95% CI due to only 1 event in the mRNA-1273 arm and the overall small sample size of Werbel 2021.

^g^Lower grading due to imprecision, higher grading due to strong association in RR. Type 3 due to nonrandomized studies.

^h^*I*²=31.3%, Χ²=17.47, p(Q)=0.13; no issues of heterogeneity or inconsistency.

^i^Wider 95% CI due to only 3 events in the mRNA-1273 arm and overall small sample size in Hammer 2022.

^j^*I*²=67.1%, Χ²=18.24, p(Q)=0.01; substantial heterogeneity.

^k^Lower grading due to indirectness and higher grading due to strong association in RR. Type 3 due to nonrandomized studies.

^m^*I*²=59.6%, Χ²=37.15, p(Q)=0; substantial heterogeneity.

^n^Higher grading due to strong association in RR. No issues with imprecision. Type 3 due to nonrandomized studies.

^o^*I*²=57.0%, Χ²=16.28, p(Q)=0.02; substantial heterogeneity.
