## Supplemental Table 10 for "Immunogenicity of mRNA-1273 and BNT162b2 in Immunocompromised Patients: Systematic Review and Meta-Analysis Using GRADE"

### Table S10. GRADE Summary of Findings for Total Anti-spike Binding Antibody or IgG Titers by Disease Subgroup and Anti-CD20 Treatment

|  | **Certainty Assessment** | | | | | | | **Relative Increase (95% CI)** | **Certainty** | **Importance** |
| --- | --- | --- | --- | --- | --- | --- | --- | --- | --- | --- |
|  | **Studies, n** | **Study Design** | **RoB** | **Inconsistency** | **Indirectness** | **Imprecision** | **Other  Considerations** |  |  |  |
| **Autoimmune disease** |  | | | | | | | | | |
| **Anti-CD20 mAb treatment** | 6 | nR | NS | Serious^a^ | Serious^b^ | Very serious^c^ | Strong association | 21.21% (3.78%, 38.63%) | Type 3^d^ | Critical |
| **No anti-CD20 mAb treatment** | 10 | nR | NS | Serious^e^ | Serious^b^ | Very serious^f^ | Strong association | 119.95% (52.01%, 187.89%) | Type 3^d^ | Critical |
| **Hematologic malignancy** |  | | | | | | | | | |
| **Anti-CD20 mAb treatment** | 5 | nR | NS | NS^g^ | Serious^b^ | Very serious^h^ | None | −12.25% (−27.83%, 3.34%) | Type 4^i^ | Critical |
| **No anti-CD20 mAb treatment** | 6 | nR | NS | Serious^j^ | Serious^b^ | NS | None | 50.82% (−13.28%, 114.92%) | Type 3^k^ | Critical |

GRADE, Grading of Recommendations, Assessment, Development, and Evaluation; IgG, immunoglobulin G; mAb, monoclonal antibody; nR, nonrandomized; NS, not serious; RoB, risk of bias.

^a^*I*²=55.2%, Χ²=11.17, p(Q)=0.05; moderate heterogeneity.

^b^Outcome definitions were rather heterogeneous (various antibody types tested through various assays).

^c^Wide 95% CI for mean antibody levels per arm in Hammer 2022.

^d^Lower grading due to imprecision, higher grading due to strong association in relative increase/decrease. Type 3 due to nonrandomized studies.

^e^*I*²=90.7%, Χ²=96.65, p(Q)=0; considerable heterogeneity.

^f^Wide 95% CI because of imputation of standard error by mean standard deviation over all studies in analysis in Verstappen 2022.

^g^*I*²=20.0%, Χ²=5.0, p(Q)=0.37; no issues of heterogeneity or inconsistency.

^h^Wide 95% CI for mean antibody levels per arm in Nishikubo 2022.

^i^Lower grading due to imprecision and nonrandomized studies. No strong association identifed, therefore type 4.

^j^*I*²=85.9%, Χ²=35.56, p(Q)=0; considerable heterogeneity.

^k^Lower grading due to no strong association identified and nonrandomized studies. No issues with imprecision identifed, therefore type 3.
