## Supplemental Table 11 for "Immunogenicity of mRNA-1273 and BNT162b2 in Immunocompromised Patients: Systematic Review and Meta-Analysis Using GRADE"

### Table S11. GRADE Summary of Findings for Neutralizing Anti-spike Titers by Disease Subgroup and Anti-CD20 Treatment

|  | **Certainty Assessment** | | | | | | | **Relative Increase (95% CI)** | **Certainty** | **Importance** |
| --- | --- | --- | --- | --- | --- | --- | --- | --- | --- | --- |
|  | **Studies, n** | **Study Design** | **RoB** | **Inconsistency** | **Indirectness** | **Imprecision** | **Other Considerations** |  |  |  |
| **Autoimmune disease** |  | | | | | | | | | |
| **Anti-CD20 mAb treatment** | 2 | nR | NS | NS^a^ | Serious^b^ | NS | Limited evidence | 28.73% (−75.57%, 133.03%) | Type 4^c^ | Limited evidence |
| **No anti-CD20 mAb treatment** | 1 | nR | NS | NS^d^ | Serious^b^ | Very serious^e^ | Limited evidence | 49.02% (−144.35%, 242.39%) | Type 4^c^ | Critical |

GRADE, Grading of Recommendations, Assessment, Development, and Evaluation; IQR, interquartile range; mAb, monoclonal antibody; nR, nonrandomized; NS, not serious; RoB, risk of bias.

^a^*I*²=12.3%, Χ²=1.14, p(Q)=0.29; no issues of heterogeneity or inconsistency.

^b^Outcome definitions were rather heterogeneous (various antibody types tested through various assays).

^c^Lower grading due limited evidence and nonrandomized studies, therefore type 4.

^d^*I*²=0%, Χ²=0, p(Q)=1; no issues of heterogeneity or inconsistency.

^e^Wide 95% CI because of wide IQR for mean antibody levels per arm in Andreica 2022.

Because only 1 study reporting neutralizing anti-spike titers in patients with hematologic malignancies was included in the meta-analysis, this subgroup was not available for the anti-CD20 subgroup analysis.
