## Supplemental Table 12 for "Immunogenicity of mRNA-1273 and BNT162b2 in Immunocompromised Patients: Systematic Review and Meta-Analysis Using GRADE"

### Table S12. GRADE Summary of Findings for Cellular Immune Response by Disease Subgroup by Anti-CD20 Treatment

|  | **Certainty Assessment** | | | | | | | **Relative Increase (95% CI)** | **Certainty** | **Importance** |
| --- | --- | --- | --- | --- | --- | --- | --- | --- | --- | --- |
|  | **Studies, n** | **Study Design** | **RoB** | **Inconsistency** | **Indirectness** | **Imprecision** | **Other Considerations** |  |  |  |
| **Autoimmune disease** |  | | | | | | | | | |
| **Anti-CD20 mAb treatment** | 2 | nR | NS | Serious^a^ | Serious^b^ | Very serious^c^ | Limited evidence | 5.89% (−42.21%, 53.99%) | Type 4^d^ | Limited evidence |
| **No anti-CD20 mAb treatment** | 4 | nR | NS | Serious^e^ | Serious^b^ | Very serious^f^ | None | 17.83% (−31.92%, 67.57%) | Type 4^g^ | Critical |
| **Hematologic malignancies** |  | | | | | | | | | |
| **Anti-CD20 mAb treatment** | 2 | nR | NS | Serious^h^ | Serious^b^ | Very serious^i^ | Limited evidence | 18.07% (−66.31%, 102.45%) | Type 4^d^ | Critical |
| **No anti-CD20 mAb treatment** | 1 | nR | NS | NS^j^ | Serious^b^ | NS | Limited evidence | 102.30% (2.62%, 201.98%) | Type 3^k^ | Limited evidence |

GRADE, Grading of Recommendations, Assessment, Development, and Evaluation; mAb, monoclonal antibody; nR, nonrandomized; NS, not serious; RoB, risk of bias.

^a^*I*²=78.2%, Χ²=4.58, p(Q)=0.03; considerable heterogeneity.

^b^Outcome definitions were rather heterogeneous (various antibody types tested through various assays).

^c^Wide 95% CI for mean antibody levels per arm in Alfonso-Dunn 2022.

^d^Lower grading due to imprecision, limited evidence, and nonrandomized studies. No strong association identifed, therefore type 4.

^e^*I*²=61.1%, Χ²=7.71, p(Q)=0.05; substantial heterogeneity.

^f^Wide 95% CI because of imputation of standard error by mean standard deviation over all studies in analysis in Verstappen 2022.

^g^Lower grading due to imprecision and nonrandomized studies. No strong association identifed, therefore type 4.

^h^*I*²=67.9%, Χ²=3.12, p(Q)=0.08; substantial heterogeneity.

^i^Wide 95% CI for mean antibody levels per arm in Mairhofer 2021.

^j^*I*²=0%, Χ²=0, p(Q)=1; no issues of heterogeneity or inconsistency.

^k^Lower grading due to limited evidence, higher grading due to strong association in relative increase or decrease. Type 3 due to nonrandomized studies.
